# Human papillomavirus knowledge and associated factors in Cameroon: a systematic review and meta-analysis

**DOI:** 10.64898/2026.04.28.26351969

**Authors:** Fabrice Zobel Lekeumo Cheuyem, Arole Darwin Touko, Chabeja Achangwa, Rick Tchamani, Eno Ethel Ambo, Bérenger Luc Thierry Bihina Noah, Constantine Tanywe Asahngwa

**Author notes:** **Corresponding author address**: Fabrice Zobel Lekeumo Cheuyem.

## Abstract

**Background:** Human papillomavirus (HPV) infection is a major public health concern in Cameroon, where cervical cancer remains the second leading cause of cancer-related morbidity and mortality among women. Despite the availability of effective preventive measures, their uptake remains suboptimal and is influenced by population-level knowledge and awareness. This study aimed to synthesize existing evidence on HPV-related knowledge and its associated factors in Cameroon.

**Methods:** This review included studies assessing knowledge of HPV as a sexually transmitted infection (STI), its causal role in cervical cancer, and overall good HPV knowledge. A comprehensive and systematic search was conducted across PubMed, Scopus, Web of Science, Embase, the Cochrane Library, and local online databases. Study quality was appraised using the Joanna Briggs Institute critical appraisal tool. Pooled prevalence estimates were calculated using random-effects models (DerSimonian and Laird). Heterogeneity was assessed using the *I*² statistic and explored through subgroup analyses.

**Results:** A total of 32 studies involving 13,□457 participants were included. The pooled prevalence of overall good HPV knowledge was 27.4% (95% CI: 7.6–63.2; 7 studies; n = 3,312), with considerable heterogeneity (*I*² = 99.3%). Knowledge of HPV as a cause of cervical cancer was 27.9% (95% CI: 15.8–44.4; 26 studies; n = 8,688), while knowledge of HPV as an STI was 47.1% (95% CI: 31.4–63.5; 18 studies; n = 9,040). Healthcare workers demonstrated the highest levels of knowledge (80.2% for HPV as an STI; 78.7% for HPV as a cause of cervical cancer), whereas students (43.4% and 10.2%, respectively) and women from the general population (30.6% and 19.9%, respectively) showed substantially lower levels. Factors associated with poor knowledge included Christian affiliation (OR = 1.46; 95% CI: 0.08–26.06) and secondary level education (OR = 1.32; 95% CI: 0.66–2.63), although these associations were non-significant.

**Conclusions:** This study reveals that, HPV-related knowledge in Cameroon remains low, particularly regarding the causal link between HPV and cervical cancer. These findings highlight the urgent need for targeted, context-specific educational interventions and strengthened public health strategies to improve awareness and uptake of HPV prevention measures.

**Systematic review registration:** PROSPERO CRD420261283152.

## Background

The Human Papillomavirus (HPV) is currently recognized as the most common existing sexually transmitted infection (STI) worldwide, including a set of viral species with a considerable potential for oncogenesis [1]. In this regard, notably high-risk viral species 16 and 18 are classified as the main causative agents of cervical cancer, accounting for around 70% of all existing cervical cancer cases [2]. Although HPVs are a concern from a global public health point of view, their burden is considerably higher in low and middle-income countries due to limitations in both primary and secondary preventive measures such as HPV vaccination, sexual health education, and cervical cancer screening [2].

The impact of the virus is most severe in Cameroon. Currently, it stands out as the second most common cancer among women, as well as the second leading cause of cancer-related mortality [3]. Statistics indicate that at least 1,500 Cameroonian women are diagnosed with cervical cancer annually, while close to 1,000 deaths occur each year. This risk is heightened by an early average age of sexual debut, estimated at about 16.7 years, therefore increasing the window for exposure to high-risk viral strains [4]. Despite the introduction of a free quadrivalent vaccine to stem its burden into the National Immunization Program in 2019, uptake remains suboptimal, with coverage currently estimated around 20% [5].

The difference between the availability of life-saving interventions and their utilization on the ground is deeply entrenched in the socio-cognitive landscape of the population [1]. The association between chronic viral carriage and the emergence of malignancy is often clouded by local myths and cultural beliefs. In most communities, the causative agent of the disease is thought to be poor hygiene, deviant behavior, or a form of medical device-induced damage, rather than a virus [1]. Even more, local misconceptions persist among some healthcare staff who are supposed to provide reliable health information. A study found that close to one-third of the nursing staff do not consider the virus to be sexually transmitted agent, and many mistakenly believe the infection is uncommon [6].

Social resistance to HPV vaccination and cervical cancer prevention efforts contributes to the complexity of the issue and spreading of the disease. Misconceptions about safety of the vaccines and its use as a means of sterilization make people become apprehensive about the issue [1]. Misconceptions prove to be much more convincing than facts. In some conflict prone rural areas where access to health education is not available to the public to the same degree, the choice to obtain the preventive method does not depend solely on distance to the health facilities but rather on the background and marital status of the person [3].

There have been several studies conducted in different geographical locations in Cameroon exploring HPV knowledge, attitudes, and acceptability of HPV vaccine in adolescents, parents, and medical practitioners [4,6–9]. These studies exhibit considerable methodological heterogeneity, assess diverse outcomes, and lack national representativeness, thereby limiting the generalizability of findings regarding HPV health literacy and its associated social determinants. This constitutes a critical gap in a country where coverage for both HPV vaccination and cervical cancer screening remains low. Based on existing literature, no systematic or scoping review is ongoing or published on this specific topic in Cameroon. This is an indication that decision-makers in policy and clinical practice currently lack a comprehensive understanding of the level of knowledge of HPV and the factors that shape this knowledge. Consequently, interventions aimed at tackling the spread of HPV have not been informed by the best available evidence synthesis, given that empirical studies on this topic had not been synthesized, despite the availability of primary studies [1,7–35]. The present systematic review and meta-analysis aim to synthesize the best available evidence on people’s knowledge of HPV and influential factors to inform better interventions aimed at improving understanding of HPV amongst community members and health care professionals.

## Method

### 1.1. Study design and protocol registration

This systematic review and meta-analysis were conducted and reported in accordance with the Preferred Reporting Items for Systematic Reviews and Meta-Analyses (PRISMA) guidelines. The study protocol was prospectively registered with the International Prospective Register of Systematic Reviews (PROSPERO) under the registration number CRD420261283152.

### 1.2. Eligibility Criteria

#### Inclusion criteria

This review included quantitative, observational studies conducted before 2025 in Cameroon among adolescents, adults, parents or caregivers, healthcare workers, vaccinators, or community members. Interventional studies that measure at least our outcome of interest were also eligible. To be eligible, the study had to report at least one of the following domains as an HPV-related knowledge: awareness of HPV as a sexually transmitted infection, knowledge of the link between HPV and cervical cancer, and overall level of HPV knowledge. Only studies written in English or French were included.

#### Exclusion criteria

Studies conducted outside Cameroon, qualitative studies, review articles, editorials, commentaries, conference abstracts, and letter to editor were excluded. Studies lacking sufficient data (full-text) to estimate these specific prevalences were also disregarded.

### Search strategy

### 1.3. Databases

A comprehensive literature search was conducted in PubMed, Scopus, the Cochrane Library, Web of Science, Embase, African Journals Online (AJOL), and Health Sciences and Diseases. Moreover, manual search was conducted on google scholar. Reference lists of included studies were also screened to identify additional eligible articles. Screening was performed independently by two investigators (FZLC and ADT), with any disagreements resolved through discussion.

### 1.4. Search terms

Our search strategy utilized Medical Subject Headings (MeSH) and free-text terms combined with Boolean operators: (“human papillomavirus” OR HPV OR “cervical cancer”) AND (knowledge OR awareness OR attitude OR practice) AND (Cameroon OR Cameroonian OR Cameroun). For the PubMed repository, the specific search string was as follows: (“human papillomavirus” OR “HPV” OR “cervical cancer”) AND (“knowledge” OR “attitude” OR “awareness” OR “practice”) AND (“Cameroon” OR “Cameroonian” OR “Cameroun”). The final search was conducted on January 27, 2025. (**Supplementary Material 1: Table 1**).

**Table 1.**
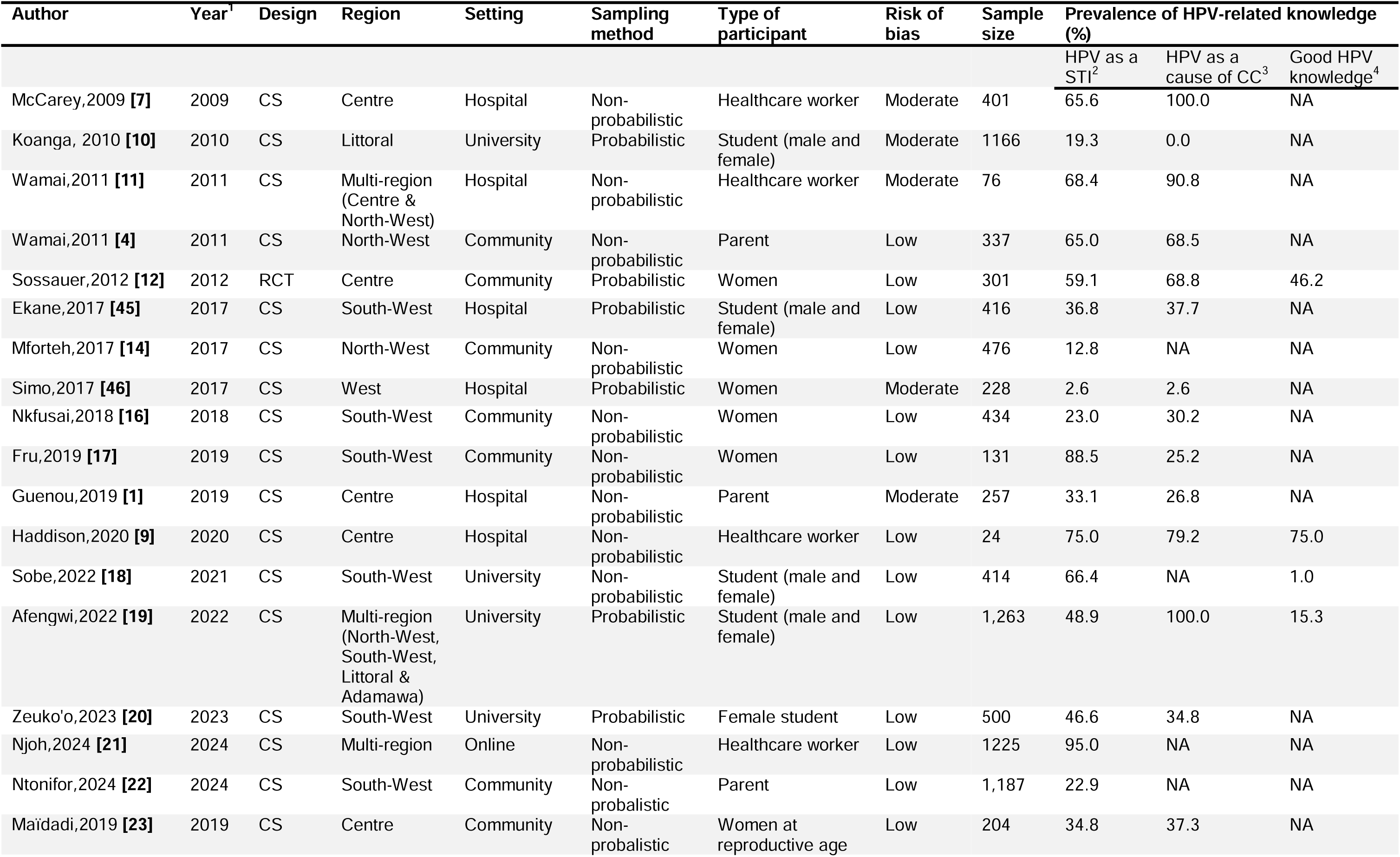
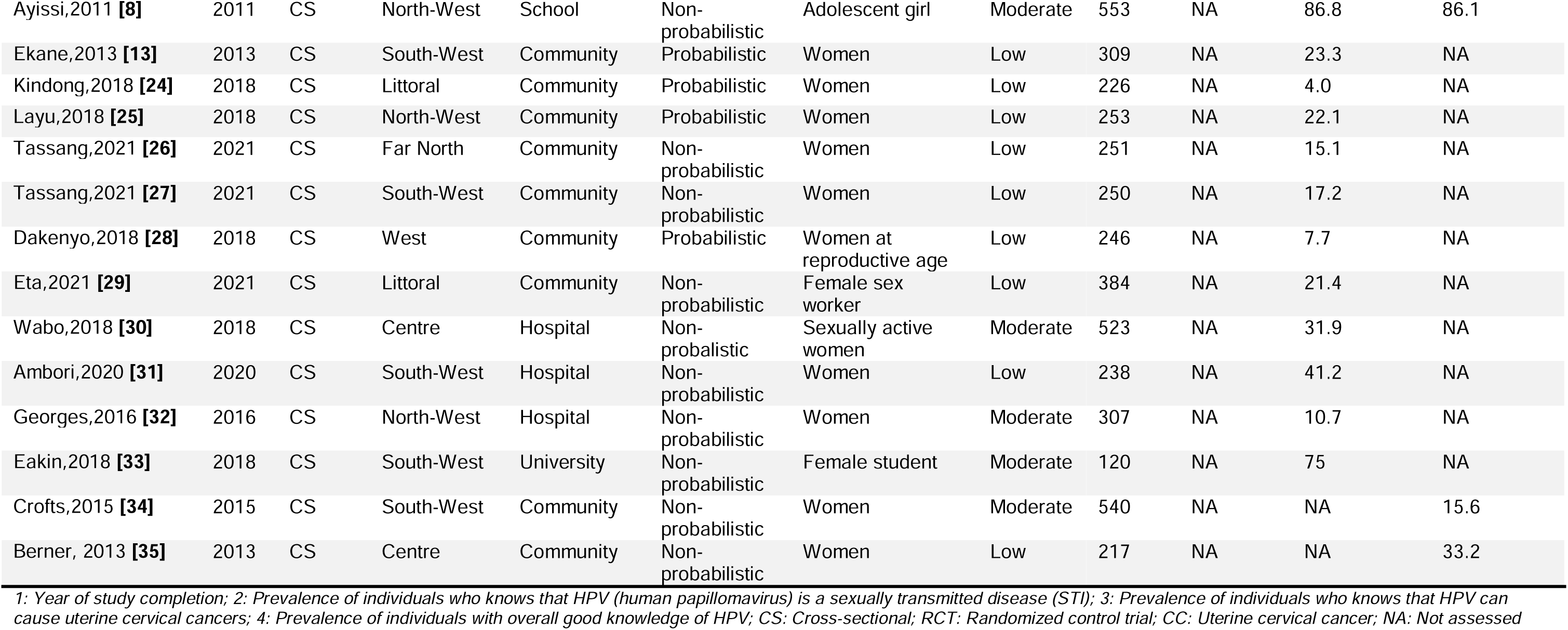
Synthesis of included study characteristics.

### 1.5. Data extraction

Data were extracted by two independent investigators (FZLC and CA) using a standardized form, with discrepancies resolved through consensus or consultation with a third reviewer (CTA). The extracted variables comprised study characteristics (author, year of completion, region, setting, and design), target population, sampling methods, and sample size. To determine pooled odds ratios, we extracted the number of individuals in both exposed and unexposed/control groups for each specific predictor of poor knowledge. The outcomes of interest included knowledge of HPV as a sexually transmitted infection, awareness of HPV as a cause of cervical cancer, and overall good HPV knowledge.

### 1.6. Outcome measurement and operational definition

Our primary outcome was the prevalence of HPV knowledge as a sexually transmitted infection (STI), defined as the proportion of individuals who correctly identified or were aware that HPV is an infection transmissible through sexual intercourse. We also assessed the prevalence of knowledge of HPV as a cause of cervical cancer, referring to the proportion of individuals who correctly identified HPV as a risk factor for, or a cause of, cervical cancer. Our secondary outcome encompassed the prevalence of overall good knowledge of HPV, representing the proportion of individuals categorized as having a sufficient level of knowledge by study investigators based on a series of questions related to HPV.

### 1.7. Data quality assessment

The risk of bias in the included articles was assessed using the Joana Briggs Institute critical appraisal checklist [36]. The number of parameters used to assess the quality of the included manuscript depended on the study design. For studies reporting prevalence data the items included : (1) sample frame appropriate to address the target population, (2) study participants sampled in an appropriate way, (3) sample size adequate, (4) study subjects and the setting described in detail, (5) data analysis conducted with sufficient coverage of the identified sample, (6) valid methods used for the identification of the condition, (7) condition measured in a standard, reliable way for all participants, (8) appropriate statistical analysis, (9) response rate adequate, and or low response rate managed appropriately. Each parameter will be scored as 1 (yes) or 0 (no or unclear). The risk of bias will be categorized as low: 7-9, moderate: 6-5, or high: 3-0 [37,38]. Two reviewers independently assess the risk of bias. Disagreement between the reviewers were resolved by discussion to reach consensus (FZLC, RT) or consultation with a third reviewer (CTA).

### 1.8. Statistical analysis and synthesis

The *I^2^* statistic was used to assess heterogeneity between studies. A DerSimonian and Laird random-effects model was chosen to pool the estimates, anticipating significant heterogeneity. Generalized linear mixed models (GLMM), coupled with the logit transformation (PLOGIT), were employed for their effectiveness in handling meta-analyses of binary data [39]. For each study outcome, prevalence estimates and corresponding 95% confidence intervals (CIs) were calculated.

To explore variability in results, subgroup analyses were performed based on study timeframe, region, population group, study setting, and sampling method. Statistical significance was defined as a *p*-value of <0.05. All analyses were conducted using the ‘meta’ package in R version 4.5.2 [40]. Additionally, predictors of “good knowledge” of HPV were analyzed to generate pooled odds ratios (ORs) and their corresponding 95% CIs.

### 1.9. Publication bias and sensitivity analysis

Publication bias was assessed visually using funnel plots and statistically through Egger’s or Begg’s tests, provided at least 10 studies were included in the meta-analysis [41,42]. A *p*-value□>□0.05 was considered indicative of no statistically significant evidence of publication bias. To evaluate the robustness of the pooled estimates, a sensitivity analysis was performed using the leave-one-out method (iteratively excluding one study at a time). Additionally, the Duval and Tweedie trim-and-fill method was employed to adjust for and further explore the impact of potential publication bias [43].

## Results

### 1.10. Studies selection

We identified 407 articles through database searches and 2,154 from additional sources. After removing 21 duplicates, we screened the remaining titles and abstracts, excluding 340 reports. Of the 96 articles assessed for eligibility, 32 were ultimately included in the systematic review and meta-analysis. Specifically, 18 articles were used to assess prevalence of HPV knowledge as an STI, 26 focused on HPV as a cause of cervical cancer, and 8 evaluated overall good knowledge levels on HPV in Cameroon (**Fig.1**).

**Fig. 1.**
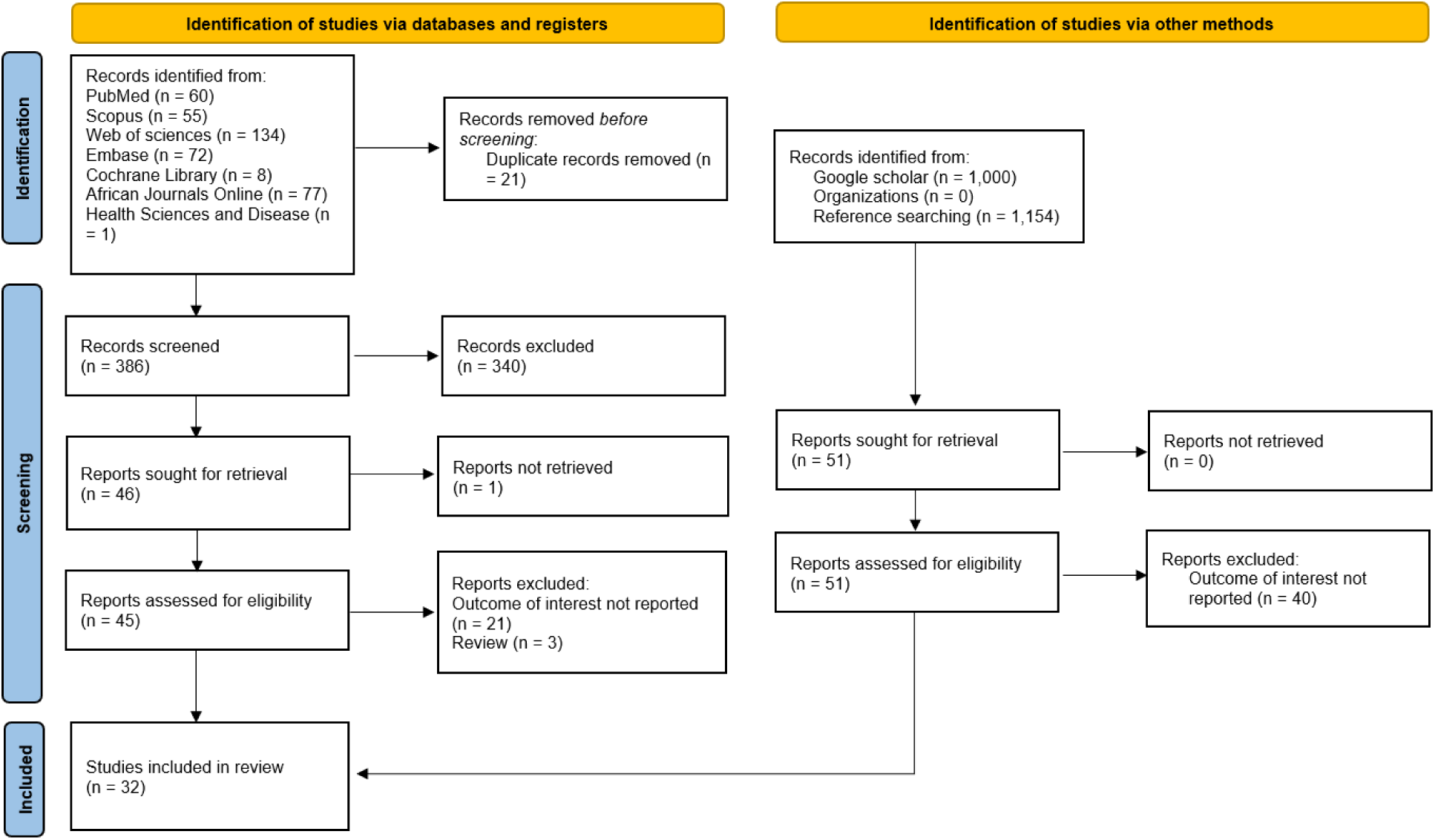
PRISMA diagram flow from study identification to inclusion in the systematic review and meta-analysis [44]

### 1.11. Study characteristics

A total of 32 study involving 13,□457 participants were included in this systematic review and meta-analysis. Most included studies were cross-sectional (n = 31; 97%) and were conducted from 2014 to 2020, corresponding to the HPV vaccination demonstration period in Cameroon (n = 16; 50%). The most frequently studied Cameroonian regions included the South-West (n = 12; 38%), followed by the Centre (n = 8; 25%), the North-West (n = 7; 22%), and the Littoral (n = 4; 13%) regions. Included reports were primarily community-based (n = 16; 50%), followed by hospital-based (n = 9; 28%), and university/school-based (n = 6; 19%). Most studies used a non-probabilistic sampling method (n = 22; 69%). The most common participant types were women from the general population (n = 16; 50%), followed by students (n = 6; 19%), healthcare workers (n = 4; 13%), and parents (n = 3; 9%). The risk of bias as low for most of study methodologies (n = 22; 69%) (**Table 1**).

### 1.1. Knowledge of human papillomavirus as a sexually transmitted infection

The pooled prevalence of knowledge of HPV as an STI was 47.1% (95% CI: 31.4–63.5; 18 studies; n = 9,040), with high heterogeneity across studies (*I*^2^ = 99.0%; *p* < 0.001). Prevalences varied from 2.6% in a hospital-based study conducted among women in the general population in the West region in 2017, to 95% in an online study conducted across multiple regions among healthcare workers in 2024 (**Fig. 2**).

**Fig. 2.**
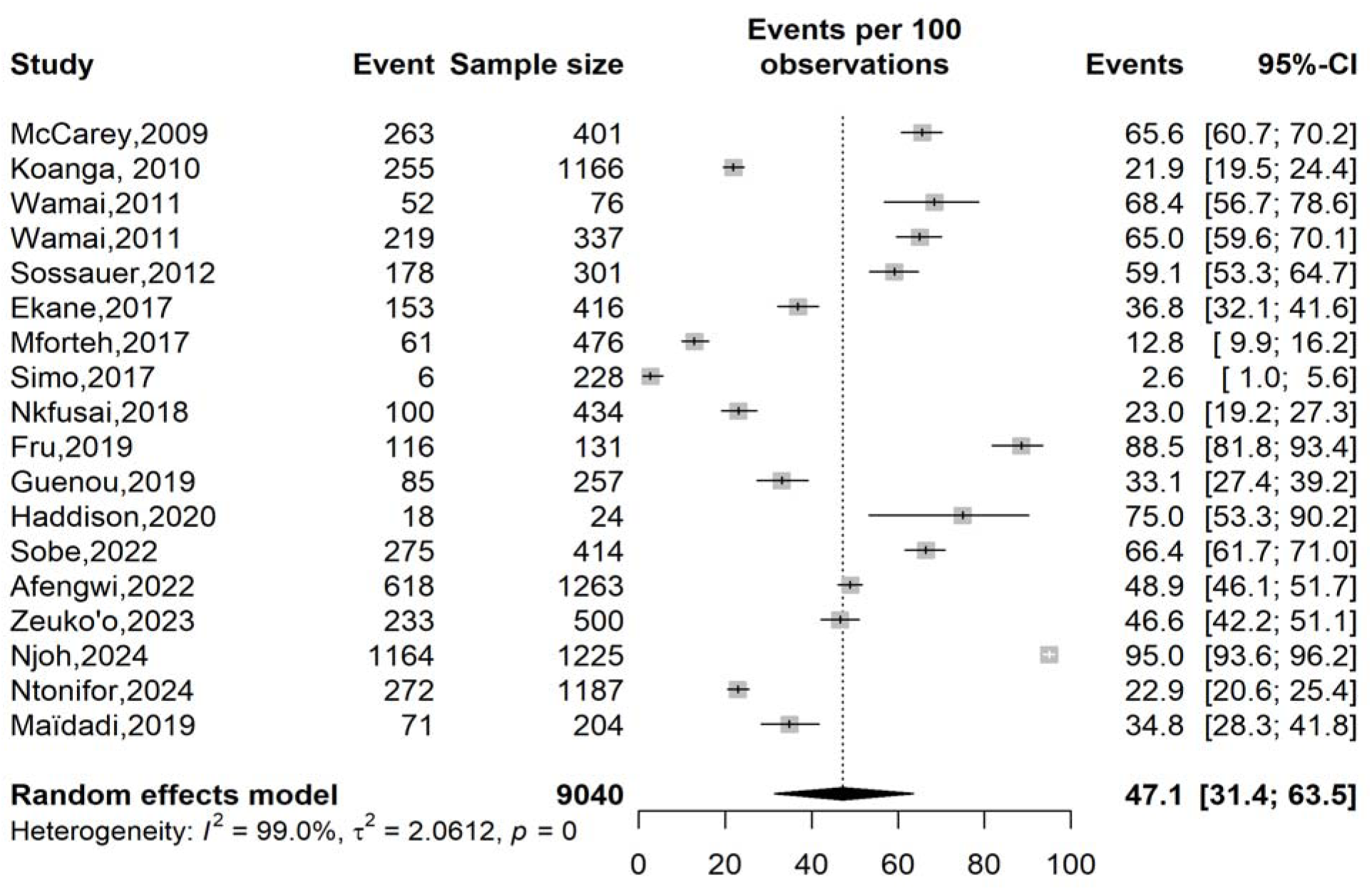
Forest plot displaying the prevalence of human papillomavirus knowledge as a sexually transmitted infection in Cameroon

The study setting (*p* < 0.001), the region of study implementation (*p* < 0.001), and the type of study participants (*p* = 0.011) were significant sources of heterogeneity between included reports. Almost similar levels of knowledge were observed among participants of studies conducted in hospitals (40.5%; 95% CI: 16.2–70.6; 6 studies; n = 1,402), in communities (42.9%; 95% CI: 22.9–65.6; 7 studies; n = 3,070), or within university institutions (45.1%; 95% CI: 29.3–62.0; 4 studies; n = 3,343).

The level of knowledge was above average in studies conducted in the Centre region (52.6 95% CI 37.8 -66.9; 5 studies; n = 1,187), and multiregional studies (77.4%; 95%CI: 44.8-93.6; 3 studies; n = 2,564) compared to study other regions where prevalence was below average.

Healthcare workers presented the highest knowledge prevalence (80.2%; 95% CI: 60.9-91.4; 4 studies, n = 1,726) compared to other groups including students (43.4%; 95%CI: 30.4-57.4; 5 studies; n = 3,759), parents of HPV vaccine eligible children (39.3%; 95%CI: 21.2-60.8; 3 studies; n = 1,781) and women from general population (30.6%; 95%CI: 9.7-64.4; 6 studies; n = 1,774) (**Table 2 and Supplementary Material 1 : Fig.1-5**).

**Table 2.** Subgroup meta-analysis of the pooled prevalence of human papillomavirus knowledge as a sexually transmitted disease in Cameroon.

| Subgroup | Sample size | Prevalence<sup>1</sup> (%) | 95% CI limits<sup>1</sup> |  | Number of studies | Heterogeneity statistic<sup>1</sup> |  | Subgroup difference |
| --- | --- | --- | --- | --- | --- | --- | --- | --- |
|  |  |  | Lower | Upper |  | I<sup>2</sup>(%) | p-value |  |
| **Study period (year)<sup>2</sup>** |  |  |  |  |  |  |  | 0.314 |
| □ 2014 | 2,281 | 55.4 | 38.6 | 71.0 | 5 | 99.0 | <0.001 |  |
| 2014-2020 | 2,170 | 33.4 | 14.1 | 60.5 | 8 | 97.3 | <0.001 |  |
| 2021-2024 | 4,589 | 61.0 | 31.6 | 84.1 | 5 | 99.5 | <0.001 |  |
| **Study setting** |  |  |  |  |  |  |  | <0.001 |
| Hospital | 1,402 | 40.5 | 16.2 | 70.6 | 6 | 97.2 | <0.001 |  |
| University | 3,343 | 45.1 | 29.3 | 62.0 | 4 | 99.0 | <0.001 |  |
| Community | 3,070 | 42.9 | 22.9 | 65.6 | 7 | 98.7 | <0.001 |  |
| Online | 1,225 | 95.0 | 93.7 | 96.2 | 1 | — | — |  |
| **Region** |  |  |  |  |  |  |  | <0.001 |
| Centre | 1,187 | 52.6 | 37.8 | 66.9 | 5 | 95.9 | <0.001 |  |
| Littoral | 1,166 | 21.9 | 19.5 | 24.4 | 1 | — | — |  |
| North-West | 813 | 34.3 | 8.2 | 75.3 | 2 | 99.5 | <0.001 |  |
| South-West | 3,082 | 48.1 | 27.3 | 69.6 | 6 | 98.7 | <0.001 |  |
| West | 228 | 2.6 | 1.0 | 5.6 | 1 | — | — |  |
| Multi-region | 2,564 | 77.4 | 44.8 | 93.6 | 3 | 99.5 | <0.001 |  |
| **Sampling method** |  |  |  |  |  |  |  | 0.095 |
| Non-probabilistic | 5,166 | 56.4 | 37.1 | 73.9 | 12 | 99.2 | <0.001 |  |
| Probabilistic | 3,874 | 30.1 | 13.3 | 54.6 | 6 | 98.4 | <0.001 |  |
| **Participant** |  |  |  |  |  |  |  | 0.011 |
| Student | 3,759 | 43.4 | 30.4 | 57.4 | 5 | 98.7 | <0.001 |  |
| Healthcare worker | 1,726 | 80.2 | 60.9 | 91.4 | 4 | 98.5 | <0.001 |  |
| Parent<sup>3</sup> | 1,781 | 39.3 | 21.2 | 60.8 | 3 | 98.9 | <0.001 |  |
| Women from general population) | 1,774 | 30.6 | 9.7 | 64.4 | 6 | 98.5 | <0.001 |  |

<sup>1</sup> Random effects model; <sup>2</sup> □ 2014 = Pre-human papillomavirus (HPV) vaccination in Cameroon, 2014-2020= HPV vaccination demonstration in Cameroon, 2021-2023= post-HPV vaccine introduction in Cameroon; <sup>3</sup> Parent of HPV vaccine eligible children

### 1.2. Knowledge of human papillomavirus as the cause of uterine cervical cancers

A low pooled estimate of HPV knowledge as a cause of uterine cervical cancer was observed among the Cameroonian population (27.9%; 95% CI: 15.8-44.4; 26 studies; n = 8,688) with high heterogeneity observed between studies (*I*^2^ = 98.2%; *p* < 0.001). The lowest prevalence (0.0%) was recorded among 1,166 participants in a study conducted among university students (male and female) in the littoral region of Cameroon), and the highest among 76 healthcare workers in a multi-regional study (Centre and North-West) (**Fig. 3**).

**Fig. 3.**
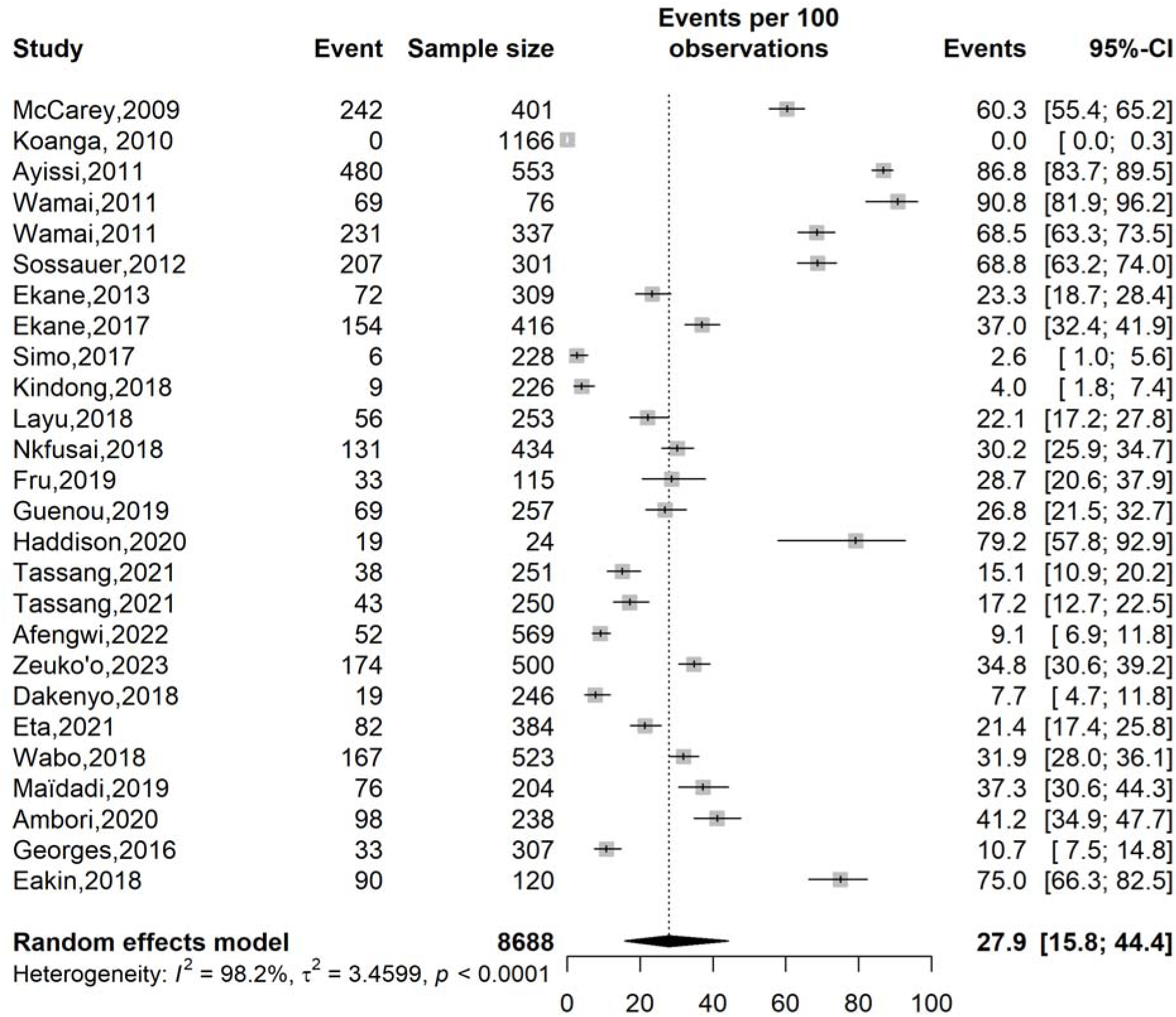
Forest plot displaying the prevalence of human papillomavirus knowledge as a cause of uterine cervical cancers in Cameroon

**Fig. 4.**
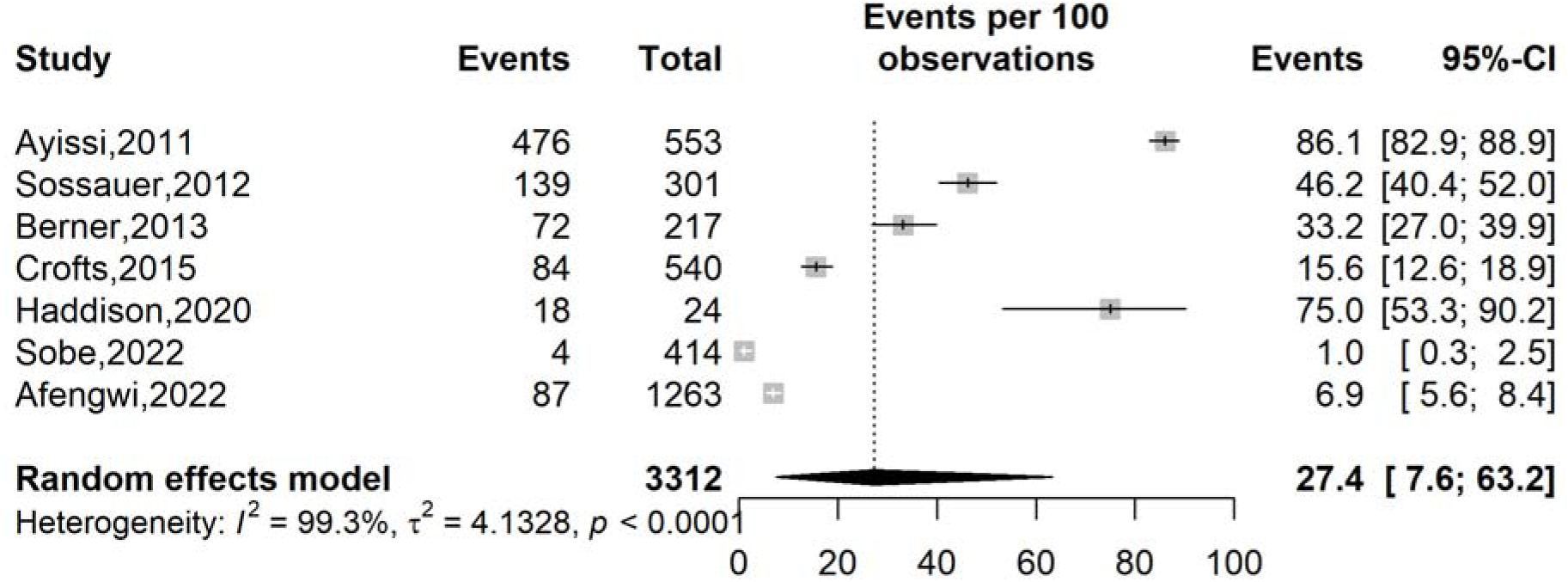
Forest plot displaying the prevalence of good knowledge on human papillomavirus in Cameroon

The study region (*p* <0.001), the sampling method (*p* = 0.010), and the type of study participants were significant source of heterogeneity across included reports. Among the pooled estimates, the highest prevalence was recorded in the Centre region (49.8%; 95% CI: 34.0–65.7; 6 studies; n = 1,710), while the lowest was observed in the West (4.8%; 95% CI: 2.2–9.9; 2 studies; n = 474) and Littoral regions (1.0%; 95% CI: 0.0–44.5; 3 studies; n = 1,776). Similar to the previous study outcome, a comparable pattern was observed for HPV knowledge as a cause of uterine cervical cancer, where healthcare workers exhibited the highest knowledge prevalence (78.7%; 95% CI: 58.7–90.6; 3 studies; n = 501) compared to other groups, including parents of HPV vaccine-eligible children (47.3%; 95% CI: 20.6–75.6; 2 studies; n = 594), women from the general population (19.9%; 95% CI: 12.2–30.7; 14 studies; n = 3,885), and students (10.2%; 95% CI: 0.5–71.6; 5 studies; n = 2,771) (**Table 3 and Supplementary Material 2 : Fig.1-5**).

**Table 3.** Subgroup meta-analysis of the pooled prevalence of human papillomavirus knowledge as a cause of uterine cervical cancers in Cameroon.

| Subgroup | Sample size | Prevalence<sup>1</sup> (%) | 95% CI limits<sup>1</sup> |  | Number of studies | Heterogeneity statistic<sup>1</sup> |  | Subgroup difference |
| --- | --- | --- | --- | --- | --- | --- | --- | --- |
|  |  |  | Lower | Upper |  | I<sup>2</sup>(%) | p-value |  |
| **Study period (year)<sup>2</sup>** |  |  |  |  |  |  |  | 0.495 |
| □ 2014 | 3,143 | 39.0 | 4.9 | 88.9 | 7 | 98.0 | <0.001 |  |
| 2014-2020 | 3,591 | 25.2 | 14.4 | 40.4 | 14 | 96.0 | <0.001 |  |
| 2021-2023 | 1,954 | 18.2 | 12.0 | 26.7 | 5 | 96.2 | <0.001 |  |
| **Study setting** |  |  |  |  |  |  |  | 0.562 |
| Community | 3,310 | 24.8 | 15.1 | 38.0 | 12 | 97.9 | <0.001 |  |
| Hospital | 2,470 | 38.0 | 17.3 | 64.1 | 9 | 97.3 | <0.001 |  |
| School and university | 2,908 | 14.7 | 0.5 | 85.5 | 5 | 99.3 | <0.001 |  |
| **Region** |  |  |  |  |  |  |  | <0.001 |
| Centre | 1,710 | 49.8 | 34.0 | 65.7 | 6 | 97.3 | <0.001 |  |
| Far North | 251 | 15.1 | 10.9 | 20.2 | 1 | — | — |  |
| Littoral | 1,776 | 1.0 | 0.0 | 44.5 | 3 | 92.6 | <0.001 |  |
| North-West | 1,450 | 45.6 | 14.9 | 80.0 | 4 | 99.3 | <0.001 |  |
| South-West | 2,382 | 34.9 | 24.5 | 46.9 | 8 | 94.4 | <0.001 |  |
| West | 474 | 4.8 | 2.2 | 9.9 | 2 | 82.1 | 0.018 |  |
| Multi-region | 645 | 45.6 | 14.9 | 80.0 | 2 | 99.2 | <0.001 |  |
| **Sampling method** |  |  |  |  |  |  |  | 0.011 |
| Non-probabilistic | 4,474 | 44.4 | 29.8 | 60.1 | 16 | 98.3 | <0.001 |  |
| Probabilistic | 4,214 | 10.1 | 2.8 | 30.7 | 10 | 98.0 | <0.001 |  |
| **Participant** |  |  |  |  |  |  |  | <0.001 |
| Adolescent girl | 553 | 86.8 | 83.7 | 89.5 | 1 | — | — |  |
| Female sex worker | 384 | 21.4 | 17.4 | 25.8 | 1 | — | — |  |
| Student | 2,771 | 10.2 | 0.5 | 71.6 | 5 | 98.0 | <0.001 |  |
| Healthcare worker | 501 | 78.7 | 58.7 | 90.6 | 3 | 91.4 | <0.001 |  |
| Parent<sup>3</sup> | 594 | 47.3 | 20.6 | 75.6 | 2 | 98.9 | <0.001 |  |
| Women from general population | 3,885 | 19.9 | 12.2 | 30.7 | 14 | 97.0 | <0.001 |  |

<sup>1</sup> Random effects model; <sup>2</sup> □ 2014 = Pre-HPV vaccination in Cameroon, 2014-2020= HPV vaccination demonstration in Cameroon, 2021-2023=

### 1.3. Good knowledge on human papillomavirus

The pooled prevalence of HPV good knowledge in Cameroon was 27.4% (95% CI: 7.6-63.2) based on 3,312 participants across 7 studies, but this estimate was influenced by high heterogeneity (*I²* = 99.3%; *p* < 0.001). There was a marked variation across studies from 1.0% to 86.1% (**Fig. 3**).

The prevalence of good knowledge was significantly influence by study period (*p* < 0.001), study setting (*p* = 0.003); study region (*p* < 0.001), and types of study participants (*p* < 0.001). The pooled estimates significantly declined with times from 58.0% (95% CI: 28.8-82.6; 3 studies; n = 1,071) before 2014 to 2.8% (95% CI: 0.7-10.9; 2 studies; n = 1,677) in 2021-2022. The lowest pooled prevalences were significantly observed within schools and university settings (14.0%; 95% CI: 0.7-78.3; 3 studies; n = 2,230), among students (2.8%; 95% CI: 0.7-10.9; 2 studies; n = 1,677) and in the south-west region (4.1%; 95% CI: 0.5-26.2; 2 studies; n **= 954) (**Table 4 and Supplementary Material 3 : Fig.1-5).

**Table 4.** Subgroup meta-analysis of the pooled prevalence of good knowledge on human papillomavirus in Cameroon.

| Subgroup | Sample size | Prevalence (%) | 95% CI limits<sup>1</sup> |  | Number of studies | Heterogeneity statistic<sup>1</sup> |  | Subgroup difference p-value |
| --- | --- | --- | --- | --- | --- | --- | --- | --- |
|  |  |  | Lower | Upper |  | I<sup>2</sup>(%) | p-value |  |
| **Study period (year)<sup>2</sup>** |  |  |  |  |  |  |  | <0.001 |
| □ 2014 | 1,071 | 58.0 | 28.8 | 82.6 | 3 | 99.1 | <0.001 |  |
| 2014-2020 | 564 | 41.4 | 9.0 | 83.5 | 2 | 97.0 | <0.001 |  |
| 2021-2022 | 1,677 | 2.8 | 0.7 | 10.9 | 2 | 93.5 | <0.001 |  |
| **Study setting** |  |  |  |  |  |  |  | 0.003 |
| Community | 1,058 | 29.9 | 17.1 | 46.9 | 3 | 97.7 | <0.001 |  |
| Hospital | 24 | 75.0 | 53.3 | 90.2 | 1 | — | — |  |
| School and University | 2,230 | 14.0 | 0.7 | 78.3 | 3 | 99.7 | <0.001 |  |
| **Region** |  |  |  |  |  |  |  | <0.001 |
| Centre | 542 | 49.2 | 30.8 | 67.8 | 3 | 88.9 | 0.001 |  |
| Multi-region | 1,263 | 6.9 | 5.6 | 8.4 | 1 | — | — |  |
| North-West | 553 | 86.1 | 82.9 | 88.9 | 1 | — | — |  |
| South-West | 954 | 4.1 | 0.5 | 26.2 | 2 | 96.9 | <0.001 |  |
| **Sampling method** |  |  |  |  |  |  |  | 0.673 |
| Non-probabilistic | 1,748 | 30.7 | 5.6 | 76.8 | 5 | 99.2 | <0.001 |  |
| Probabilistic | 1,564 | 20.1 | 4.4 | 57.9 | 2 | 99.6 | <0.001 |  |
| **Participant** |  |  |  |  |  |  |  | <0.001 |
| Adolescent girl | 553 | 86.1 | 82.9 | 88.9 | 1 | — | — |  |
| Healthcare worker | 24 | 75.0 | 53.3 | 90.2 | 1 | — | — |  |
| Student | 1,677 | 2.8 | 0.7 | 10.9 | 2 | 93.5 | <0.001 |  |
| Women from general population) | 1,058 | 29.9 | 17.1 | 46.9 | 3 | 97.7 | <0.001 |  |

<sup>1</sup> Random effects model; <sup>2</sup> □ 2014 = Pre-HPV vaccination in Cameroon, 2014-2020= HPV vaccination demonstration in Cameroon, 2021-2023= post-HPV vaccine introduction in Cameroon; <sup>3</sup> Parent of HPV vaccine eligible children

### 1.4. Predictor of poor knowledge on human papillomavirus

Although not statistically significant, younger participants (<20 years) were 20% less likely to have poor knowledge of HPV compared to their older counterparts (OR = 0.8; 95% CI: 0.4–1.4; 4 studies). Female individuals were 30% less likely to have poor knowledge of HPV compared to male individuals (OR = 0.7; 95% CI: 0.5–1.1; 2 studies). Those who identified as Christian were 46% more likely to have poor knowledge of HPV compared to individuals belonging to other faiths (OR = 1.46; 95% CI: 0.08–26.06; 3 studies). Those with a secondary school education were 32% more likely to have poor knowledge compared to those with a primary education (OR = 1.32; 95% CI: 0.66–2.63; 2 studies) (**Table 5**).

**Table 5.** Determinants of human papillomavirus poor knowledge in Cameroon.

| Predictor | Modality | OR<sup>1</sup> | 95% CI |  | Reports included | Heterogeneity |  |
| --- | --- | --- | --- | --- | --- | --- | --- |
|  |  |  | Lower | Upper |  | I<sup>2</sup>(%) | p-value |
| **Age (years)** | □ 20 vs. 20 + years | 0.8 | 0.40 | 1.40 | 4 | 83.8 | <0.001 |
| **Gender** | Female vs. Male | 0.7 | 0.50 | 1.10 | 2 | 13.1 | 0.283 |
| **Marital status** | Single vs. In partnership | 0.9 | 0.50 | 1.50 | 3 | 0.0 | 0.561 |
| **Religion** | Christian vs. Others | 1.46 | 0.08 | 26.06 | 3 | 79.2 | 0.028 |
|  | Muslim vs. Others | 0.76 | 0.31 | 1.92 | 3 | 49.3 | 0.139 |
| **Educational level** | Tertiary vs. Lower levels | 1.11 | 0.25 | 4.97 | 2 | 95.1 | <0.001 |
|  | Secondary vs. Primary | 1.32 | 0.66 | 2.63 | 2 | 2.6 | 0.311 |

<sup>†</sup>Random effect model;

### 1.5. Publication bias and sensitivity analysis

The funnel plots assessing publication bias for the pooled estimates of the three outcomes presented very slight asymmetry visually. However, the Egger’s and Begg’s test as well as the trim-and-fill analysis did not identify any significant sign of publication bias for the three outcomes (**Supplementary Fig. 7 for Supplementary Material 1-3**).

The sensitivity analysis showed no study significantly influence the pooled estimates suggesting the robustness of our findings (**Supplementary Fig. 6 for Supplementary Material 1-3**).

## Discussions

This review shows that HPV-related knowledge in Cameroon is still too low, especially when it comes to understanding that HPV is the main cause of cervical cancer. In this meta-analysis, fewer than half of participants knew that HPV is a sexually transmitted infection (47.1%), while only about one in four knew that HPV causes cervical cancer (27.9%). Overall, “good knowledge” was also low at 27.4%, and all three outcomes showed very high heterogeneity. Taken together, these findings suggest that awareness of HPV may exist at a superficial level in some groups, but deeper understanding remains limited and uneven. This trend is concerning because knowledge of the disease and its related risks is often the first step toward accepting vaccines, participating in screening and other preventive measures, and ultimately adopting appropriate behavior to prevent the disease [47,48]. For example, if a parent or adolescent has heard of HPV but does not know it is linked to cervical cancer, they may not see the vaccine as important. Our findings are consistent with recent studies from Cameroon. For instance, in Buea, a study reported that low awareness of HPV and cervical cancer was closely linked to parents hesitancy towards the vaccine uptake, along with worries about safety, side effects, distrust of health authorities and pharmaceutical companies, and limited support from religious leaders [22]. Another study from the West region found that although people generally accepted the HPV vaccine in Cameroon, knowledge and communication gaps made it difficult to implement the program [1].

Healthcare workers had the most knowledge, while students, parents, and women in the general population knew much less. This gap shows that access to information remains highly unequal, with knowledge largely concentrated among those already linked to the health system. Recent national studies support this idea. In a 2026 a survey found that health workers’ knowledge was key to improving HPV vaccine delivery and coverage, and that parents, teachers, and traditional leaders played important roles in encouraging uptake [21]. Likewise, a 2025 report from the a Cameroon Health District showed that a successful school-based HPV vaccination campaign relied on coordinated efforts by health workers, teachers, and Catholic priests, especially where rumors about infertility and COVID-19 had increased hesitancy [9]. These recent studies reinforce the main point: knowledge is important, but having trusted messengers is just as crucial [9,21].

The findings regarding time should be viewed with caution. While more people recently knew that HPV is an STI compared to earlier years, knowledge that HPV causes cervical cancer stayed low, and the percentage with “good knowledge” seemed to drop over time. This could mean that some campaigns have made people more familiar with the name HPV or the vaccine but have not clearly explained its link to cervical cancer. Also, this trend might partly be due to differences in study methods, such as who was studied and how variable the good knowledge was defined across studies. Because of this, the apparent decline should be discussed carefully and not seen as a definite national trend.

Comparing these findings in the wider sub-Saharan African context, they seem to be part of a larger regional pattern rather than just a national issue. A 2024 systematic review and meta-analysis found that HPV vaccine uptake in sub-Saharan Africa remained low overall at 29%, and that good knowledge and positive attitudes were important factors for vaccination [49]. Another meta-analysis conducted in 2025 found that parents’ willingness to vaccinate their daughters was strongly associated with good knowledge, positive attitudes, higher education, and better socioeconomic status [50]. A scoping review implemented in 2025 also found that vaccine uptake depends not only on awareness, but also on trust, gender norms, religion, school involvement, and how the vaccine is delivered [51]. These regional findings corroborate the current review and suggest that while improving knowledge helps, it is not enough unless communication is trusted and adapted to local needs.

The analysis of what predicts poor knowledge in this review should be interpreted with care. Some odds ratios suggested that younger people and females might have lower odds of poor knowledge, while those with a Christian affiliation or secondary education might have higher odds. However, none of these links were statistically significant. Some estimates, especially for religion, were also very imprecise. This means the analysis is more exploratory than confirmatory and should be interpreted with caution.

From a public health perspective, the message is clear. To improve HPV vaccine acceptance and maintain high coverage in Cameroon, communication needs to be more than just one-time awareness campaigns. Messages should explain in simple, culturally relevant ways that HPV is common, sexually transmitted, and preventable, and that vaccination against this disease can protects women against cervical cancer. Parents, adolescents, teachers, religious leaders, and healthcare workers all need to be involved. Recent experience in Cameroon shows that school- and community-based approaches work well when trusted local people participate [9,21].

In summary, this review adds to the evidence that HPV knowledge in Cameroon remains low, uneven, and closely linked to the social context in which people obtain information. Research from across sub-Saharan Africa shows that this is a common challenge [49–51] The main lesson is that improving coverage will require not just vaccine supply but also ongoing education, trusted communication, and strong community involvement.

Heterogeneity was consistently high across all main outcomes, indicating that the pooled estimates do not reflect a single, stable national level of knowledge. Rather, the findings reveal a heterogeneous landscape shaped by geographic region, study setting, participant characteristics, and likely by differences in how knowledge was measured. This variation is plausible within the Cameroonian context, where exposure to health information differs substantially across urban and rural areas, regions with varying levels of programmatic activity, and populations with differential access to and engagement with health services. This variation is not just a statistical issue; it is central to the story. It shows that HPV knowledge in Cameroon is uneven, with some groups better informed and others still left out.

### Limitations

This study has some limitations to consider. The high heterogeneity across studies may limit the generalizability of the findings. In addition, most included studies were conducted in a limited number of regions, which may not fully represent Cameroon’s national situation. Variations in how HPV good knowledge was measured across studies and the predominance of cross-sectional designs further limit comparability and causal interpretation. The analysis of predictors of poor knowledge was also inconclusive due to imprecise and non-significant estimates.

## Conclusions

In conclusion, this study demonstrates that HPV knowledge in Cameroon remains low and unevenly distributed, particularly regarding its role in cervical cancer. Despite ongoing vaccine-related communication efforts, significant gaps in awareness persist, which may hinder the success of prevention programs. Addressing these gaps through targeted, culturally appropriate education and community engagement will be essential to improving HPV vaccine uptake and reducing the burden of cervical cancer in Cameroon.

## Abbreviations

AJOL: African Journals Online
CC: Uterine cervical cancer
CI: Confidence intervals
CS: Cross-sectional
GLMM: Generalized linear mixed models
HPV: Human papillomavirus
JBI: Joanna Briggs Institute
MeSH: Medical subject headings
NA: Not assessed
OR: Odds ratio
PLOGIT: Logit transformation
PRISMA: Preferred reporting items for systematic reviews and meta-analyses
PROSPERO: International prospective register of systematic reviews
RCT: Randomized controlled trial
STI: Sexually transmitted infection

## Acknowledgements

None.

## Author contributions

Conceptualization: FZLC; Data curation: FZLC, CA, and CTA; Formal analysis: FZLC; Funding acquisition: None; Investigation: FZLC, ADT, CA, RT, and CTA; Methodology: FZLC, ADT, CA, RT, and CTA; Project administration: FZLC; Resources: All authors; Software: FZLC, ADT, CA, RT, and CTA; Supervision: FZLC and CTA; Validation: FZLC; Visualization: FZLC, ADT, CA, RT, EEA, BLTBN and CTA; Writing – original draft: FZLC, ADT, CA, and RT; Writing – review and editing: FZLC, ADT, CA, RT, EEA, BLTBN and CTA.

## Data availability

All data generated or analyzed during this study are included in this published article and its supplementary materials.

## Declarations

## Ethics approval and consent to participate

Not applicable.

## Consent to publish

Not applicable.

## Clinical trial number

Not applicable.

## References

1. Guenou E, Wakam Nkontchou B, Vouking Zambou M, Buh Nkum C, Mfoulou Minso AC, Napa YL, et al. Acceptability and Feasibility of Human Papillomavirus Vaccine Introduction in Cameroon: A Mixed-Methods Study. Cureus. 2024;16:e60723. 10.7759/cureus.60723

2. Anni NS, Rehman N, Nyambi A, Musiwa A, Graham T, Dine RD, et al. Knowledge, attitudes, and practices towards Human Papilloma Virus and uptake of HPV vaccine: A protocol for a systematic review. Karimi-Sari H, editor. PLOS ONE. 2024;19:e0313887. 10.1371/journal.pone.0313887

3. Mapoko BSE, Bell ED, Batoum V, Okobalemba EA, Fossa LT, Djampou E, et al. Knowledge, Attitudes, and Practices of University Students in a Cameroonian State University Regarding Cervical Cancer. J Cancer Ther. 2025;16:394–401. 10.4236/jct.2025.1610029

4. Wamai RG, Ayissi CA, Oduwo GO, Perlman S, Welty E, Manga S, et al. Assessing the Effectiveness of a Community-Based Sensitization Strategy in Creating Awareness About HPV, Cervical Cancer and HPV Vaccine Among Parents in North West Cameroon. J Community Health. 2012;37:917–26. 10.1007/s10900-012-9540-5

5. Njoh AA, Waheed D-E-N, Kedakse TSNJ, Ebongue LJ, Kongnyuy EJ, Amani A, et al. Overcoming challenges and achieving high HPV vaccination uptake in Cameroon: lessons learned from a gender-neutral and single-dose program and community engagement. BMC Public Health. 2025;25:1696. 10.1186/s12889-025-22776-3

6. Elit L, Ngalla C, Afugchwi GM, Tum E, Domgue JF, Nouvet E. Assessing knowledge, attitudes and belief toward HPV vaccination of parents with children aged 9–14 years in rural communities of Northwest Cameroon: a qualitative study. BMJ Open. 2022;12:e068212. 10.1136/bmjopen-2022-068212

7. McCarey C, Pirek D, Tebeu PM, Boulvain M, Doh AS, Petignat P. Awareness of HPV and cervical cancer prevention among Cameroonian healthcare workers. BMC Womens Health. 2011;11:45. 10.1186/1472-6874-11-45

8. Ayissi CA, Wamai RG, Oduwo GO, Perlman S, Welty E, Welty T, et al. Awareness, Acceptability and Uptake of Human Papilloma Virus Vaccine Among Cameroonian School-Attending Female Adolescents. J Community Health. 2012;37:1127–35. 10.1007/s10900-012-9554-z

9. Haddison E, Tambasho A, Kouamen G, Ngwafor R. Vaccinators’ Perception of HPV Vaccination in the Saa Health District of Cameroon. Front Public Health. 2022;9:748910. 10.3389/fpubh.2021.748910

10. Koanga MM, Ngono NA, Wandjis A, Dongang NR, Donfack TH, Nganwa G, et al. Sexual behaviour: human papilloma virus and cervical cancer risk among university students in cameroon. Afr J Haematol Oncol. 2010;1:115–21.

11. Wamai RG, Ayissi CA, Oduwo GO, Perlman S, Welty E, Welty T, et al. Awareness, knowledge and beliefs about HPV, cervical cancer and HPV vaccines among nurses in Cameroon: An exploratory study. Int J Nurs Stud. 2013;50:1399–406. 10.1016/j.ijnurstu.2012.12.020

12. Sossauer G, Zbinden M, Tebeu P-M, Fosso GK, Untiet S, Vassilakos P, et al. Impact of an Educational Intervention on Women’s Knowledge and Acceptability of Human Papillomavirus Self-Sampling: A Randomized Controlled Trial in Cameroon. PLOS ONE. Public Library of Science; 2014;9:e109788. 10.1371/journal.pone.0109788

13. Ekane GEH, Obinchemti TE, Nguefack CT, Nkambfu DM, Tchounzou R, Nsagha D, et al. Pap Smear Screening, the Way Forward for Prevention of Cervical Cancer? A Community Based Study in the Buea Health District, Cameroon. Open J Obstet Gynecol. 2015;05:226–33. 10.4236/ojog.2015.54033

14. Mforteh AAA, Julius DS, Merlin B, Ako TW, Walters PD, Tiku EC, et al. Factors Associated with Cervical Cancer Screening and Prevalence of Premalignant Cervical Lesions in a Rural Setting in Cameroon: a Cross-Sectional Analytical Study: Cervical cancer screening and prevalence of premalignant lesions. Health Res Afr. 2023;1:32–8. 10.5281/hra.v1i4(Suppl%201).4992

15. Tarkang E, Dzah SM, Adu-Poku F, Nzegge MM, Dadah E, Ahiabor SY. Sexual Behavior and its Association with Knowledge Regarding HIV or AIDS Among Migrant Road Construction Workers in a Rural Setting of Cameroon. J Prev Med Health Care. JSciMed Central; 2017;1:1012. 10.47739/2576-0084/1012

16. Nkfusai NC, Cumber SN, Anchang-Kimbi JK, Nji KE, Shirinde J, Anong ND. Assessment of the current state of knowledge and risk factors of cervical cancer among women in the Buea Health District, Cameroon. Pan Afr Med J. 2019 [cited 2026 Feb 17];33. 10.11604/pamj.2019.33.38.16767

17. Fru CN, Andrew T, Cho FN, Thierry T, Fru PN. Determinants of Awareness and Knowledge on Cervical Cancer among Women in Buea- Cameroon. Int J Res Rep Gynaecol. 2020;3:89–102.

18. Sobe LE. Knowledge, Attitudes and Practices Towards Human Papillomavirus (HPV) and HPV Vaccination among Students at Higher Institutions of Learning in Buea, South West Region of Cameroon: A Cross-Sectional Study. Rev. 2025;doi: 10.21203/rs.3.rs-5833643/v1. https://doi.org/10.21203/rs.3.rs-5833643/v1

19. Afegenwi MH, Feduma AS, Emilio BB, Notang MI. Human Papilloma Virus (HPV) Infection and Vaccination: Knowledge, Attitudes and Practices among Young Adults Attending Universities in Cameroon. J Adv Microbiol. 2023;23:1–14. 10.9734/jamb/2023/v23i4716

20. Zeuko’o EM, Tendem A, Ngounou E, Egbe TO. Knowledge of cervical cancer, associated risk factors, and practice of cervical cancer screening among female students in higher education institutions in Buea, Cameroon, a cross-sectional study. Clin Epidemiol Glob Health. 2025;36:102173. 10.1016/j.cegh.2025.102173

21. Njoh AA, Bolio A, Venczel L, Libwea JN, Waheed D-N, Kongnyuy EJ, et al. Key players and determinants improving human papillomavirus vaccination coverage in Cameroon: a cross-sectional nationwide health workers survey. Vaccine. 2026;69:128006. 10.1016/j.vaccine.2025.128006

22. Ntonifor MM-N, Tazinkeng NN, Kemah B-L, Claudia NE, Sonia YK, Nchinjoh SC, et al. Factors associated with parental hesitancy towards the human papillomavirus vaccine: a cross-sectional study. Sci Rep. 2025;15:18284. 10.1038/s41598-025-94067-1

23. Maïdadi FM, Bouda A, Akwa L, Mbinyui MS, Mohamadou M, Engowei MC, et al. Knowledge, Attitudes And Perception Of Women Regarding Cervical Cancer In The City Of Yaoundé, Cameroon. Br J Med Health Sci. British Journal of Medical & Health Sciences; 2020;2:203–10.

24. Kindong N-M, Akom N, Nkwain T, Nyuykighan C, Sama O, Kennedy A. Perception on Cervical Cancer, Screening Practices and its Determinants AmongWomen Aged 15-49 in Cibec and Bloc Banen Health Areas, Bonaberi Health District, Douala. Gynecol Women’s Health Care. 2025;6:1–12. 10.47485/2766-5879.1022

25. Layu D, Kifu NF, Dohbit SJ, Nkfusai CN, Bede F, Shirinde J, et al. Assessing the uptake of cervical cancer screening among women aged 25-65 years in Kumbo West Health District, Cameroon. Pan Afr Med J. 2019;33. 10.11604/pamj.2019.33.106.16975

26. Tassang A, Celestina NF, Dadao F, Domkao P, Tchounzou R, Akintunde TY, et al. Cervical cancer screening in Doukoula- the far north region of Cameroon. World J Adv Res Rev. 2023;20:404–11. 10.30574/wjarr.2023.20.2.2033

27. Tassang A, Fru CN, Brady MR, Cho FN, Thierry T, Paulette NF, et al. Knowledge and Risk Factors of Cervical Cancer among Women in Towns of Fako Division- Cameroon. J Adv Med Med Res. 2021;81–91. 10.9734/jammr/2021/v33i1631004

28. Dakenyo RD, Kenfack B, Vogue N, Tsakoue EF, Ebode ME, Cumber SN. Connaissances, attitudes et pratiques des femmes en âge de procréer du District de Santé de la Mifi sur la prévention du cancer du col de l’utérus, Cameroun. Pan Afr Med J. 2018;31. 10.11604/pamj.2018.31.172.16320

29. Eta V, Mh S, Njc A, Ds N. Knowledge, Attitudes and Practices towards Cervical Cancer Prevention among Female Sex Workers in the Douala Municipality, Cameroon. Int J Res Biol Sci. 2024;1:47–62.

30. Wabo B, Nsagha DS, Nana T, Pokam BDT, Njiomenie GF, Guemdjom WP, et al. Knowledge on cervical cancer and screening tests among women at two reference hospitals in Yaounde, Cameroon. Int J Biol Chem Sci. 2019;13:1487–95. 10.4314/ijbcs.v13i3.22

31. Ambori AC, Peter T, Ngounou E, Elombo S, Ndaka W, Tanue EA, et al. Knowledge on Cervical Cancer and its Prevention Amongst Females Attending Limbe Regional Hospital in the South-West Region of Cameroon. Int J Res Sci Innov. 2025;12:2060–70. 10.51244/IJRSI.2025.1210000182

32. Georges K, Herve AN, Simo RT, Jeremie M, Charlette N. Knowledge and behavior of women on cervical cancer in the northern region of Cameroon. Front Women’s Health. 2017;2:1–4. 10.15761/FWH.1000131

33. Eakin C, Ekollo R, Nembulefack D, Halle-Ekane G, Tangui G, Brady R, et al. Cervical Cancer Screening Beliefs and Prevalence of LSIL/HSIL Among a University-Based Population in Cameroon. J Low Genit Tract Dis. 2018;22:274–9. 10.1097/LGT.0000000000000433

34. Crofts V, Flahault E, Tebeu PM, Untiet S, Boulvain M, Vassilakos P, et al. Education efforts may contribute to wider acceptance of human papillomavirus self-sampling. Int J Womens Health. 2015;7:149–54. 10.2147/IJWH.S56307

35. Berner A, Hassel SB, Tebeu P-M, Untiet S, Kengne-Fosso G, Navarria I, et al. Human Papillomavirus Self-Sampling in Cameroon: Women’s Uncertainties Over the Reliability of the Method Are Barriers to Acceptance. J Low Genit Tract Dis. 2013;17:235–41. 10.1097/LGT.0b013e31826b7b51

36. JBI Critical Appraisal Tools. JBI. 2026. https://jbi.global/critical-appraisal-tools. Accessed 29 Mar 2026

37. Nwaozuru U, Obiezu-Umeh C, Uzodufa SA, Salako A, Akinsolu FT, Ezechi OC, et al. Systematic review and meta-analysis of the prevalence of oral cancer in Nigeria. BMC Oral Health. 2025;25:414. 10.1186/s12903-025-05724-w

38. Abraham G, Gelana B, Yitbarek K, Morankar S. Prevalence of domestic violence in a time of catastrophic disease outbreaks including COVID-19 pandemic: a systematic review protocol. Syst Rev. 2022;11:46. 10.1186/s13643-022-01920-9

39. Stijnen T, Hamza TH, Ozdemir P. Random effects meta-analysis of event outcome in the framework of the generalized linear mixed model with applications in sparse data. Stat Med. 2010;29:3046–67. 10.1002/sim.4040

40. R Core Team. R: A Language and Environment for Statistical Computing. R Found. Stat. Comput. Austria. 2024. https://www.R-project.org/. Accessed 30 May 2024

41. Egger M, Davey Smith G, Schneider M, Minder C. Bias in meta-analysis detected by a simple, graphical test. BMJ.1997;315:629–34. 10.1136/bmj.315.7109.629

42. Begg CB, Mazumdar M. Operating characteristics of a rank correlation test for publication bias. Biometrics. 1994;50:1088.

43. Duval S, Tweedie R. Trim and Fill: A Simple Funnel-Plot–Based Method of Testing and Adjusting for Publication Bias in Meta-Analysis. Biometrics. 2000;56:455–63. 10.1111/j.0006-341X.2000.00455.x

44. Page MJ, McKenzie JE, Bossuyt PM, Boutron I, Hoffmann TC, Mulrow CD, et al. The PRISMA 2020 statement: an updated guideline for reporting systematic reviews. BMJ. British Medical Journal Publishing Group; 2021;372:n71. 10.1136/bmj.n71

45. Halle-Ekane GE, Nembulefack DK, Orock GE, Fon PN, Tazinya AA, Tebeu PM. Knowledge of Cervical Cancer and Its Risk Factors, Attitudes and Practices towards Pap Smear Screening among Students in the University of Buea, Cameroon. J Cancer Tumor Int. 2018;7:1–11. 10.9734/JCTI/2018/43965

46. Tagne Simo R, Christian Kamnang T, Léonie Marthe Ghomsi S, Jacqueline F, Armel Hervé Nwabo K, Paul F Seke E, et al. Primary and Secondary Prevention of Cervical Cancer at Two District Health Centres in the West Region of Cameroon. Int J Cancer Clin Res. 2020;7:1–7. 10.23937/2378-3419/1410136

47. Alghalyini B, Zaidi ARZ, Meo SA, Faroog Z, Rashid M, Alyousef SS, et al. Awareness and knowledge of human papillomavirus, vaccine acceptability and cervical cancer among college students in Saudi Arabia. Hum Vaccines Immunother. 2024;20:2403844. 10.1080/21645515.2024.2403844

48. Al-leimon A, Al-leimon O, Abdulhaq B, Al-salieby F, Jaber A-R, Saadeh M, et al. From awareness to action: Unveiling knowledge, attitudes and testing strategies to enhance human papillomavirus vaccination uptake in Jordan. J Virus Erad. 2024;10:100380. 10.1016/j.jve.2024.100380

49. Asgedom YS, Kebede TM, Seifu BL, Mare KU, Asmare ZA, Asebe HA, et al. Human papillomavirus vaccination uptake and determinant factors among adolescent schoolgirls in sub-Saharan Africa: A systematic review and meta-analysis. Hum Vaccines Immunother. 2024;20:2326295. 10.1080/21645515.2024.2326295

50. Asmelash D, Zewdia WF, Abebe GF, Girma D, Mohammed AH, Asres A. Prevalence and predictors of parental willingness to vaccinate daughters against human papillomavirus in Sub-Saharan Africa: a systematic review and meta-analysis. Front Public Health. 2025;13:1486262. 10.3389/fpubh.2025.1486262

51. Kailemia PN, Mukami V. Exploring intersectional determinants of, and interventions for, low uptake of human papillomavirus vaccine in sub-Saharan Africa: a scoping review protocol. BMJ Open. 2025;15:e083848. 10.1136/bmjopen-2023-083848

