## Supplementary Material 2 for "Human papillomavirus knowledge and associated factors in Cameroon: a systematic review and meta-analysis"

**Subgroup analysis**

**Study period**

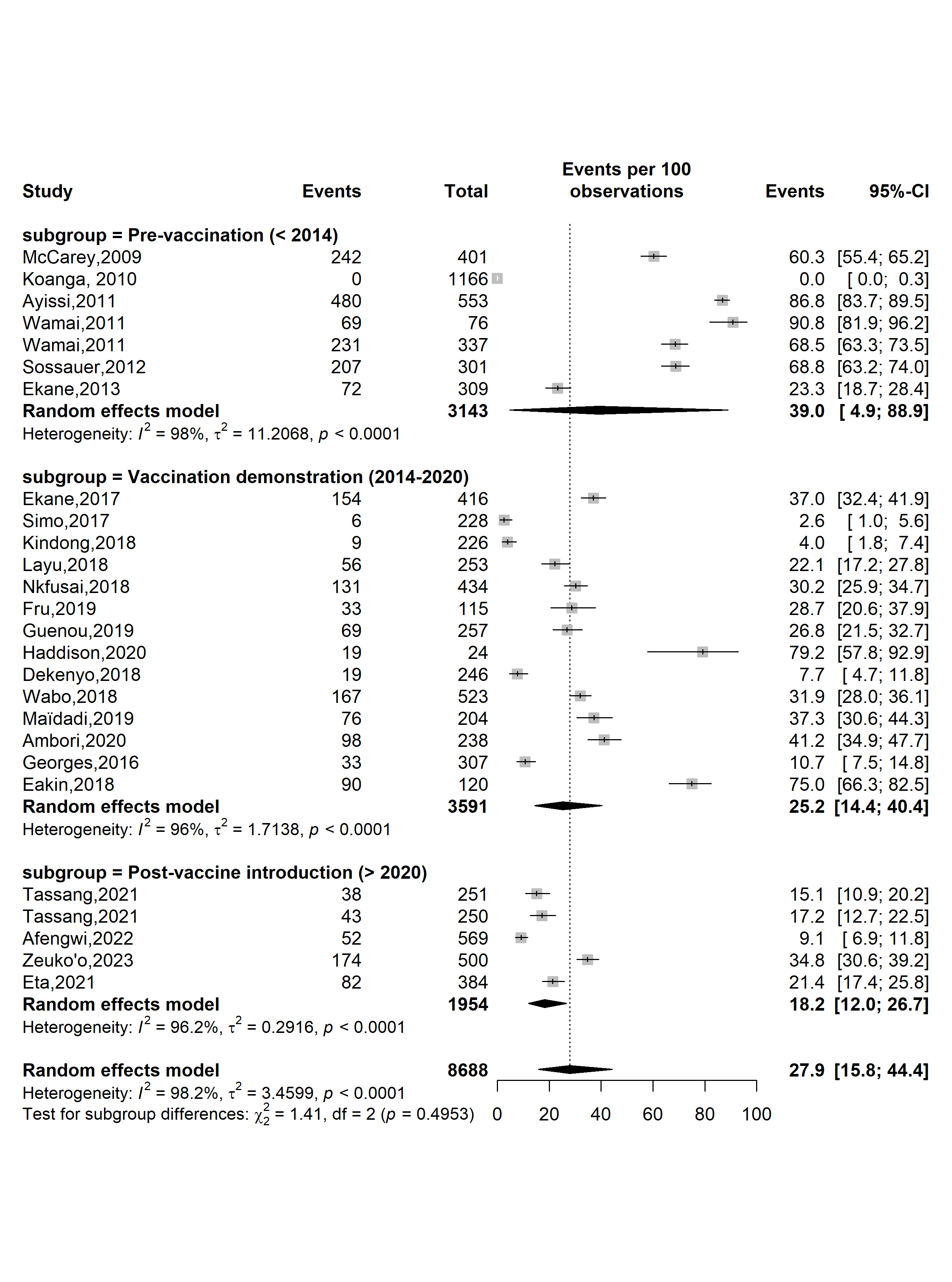

**Supplementary Fig. 1** Pooled prevalence of human papillomavirus (HPV) knowledge as a cause of uterine cervical cancers by HPV vaccine introduction timeframe in Cameroon

**Study setting**

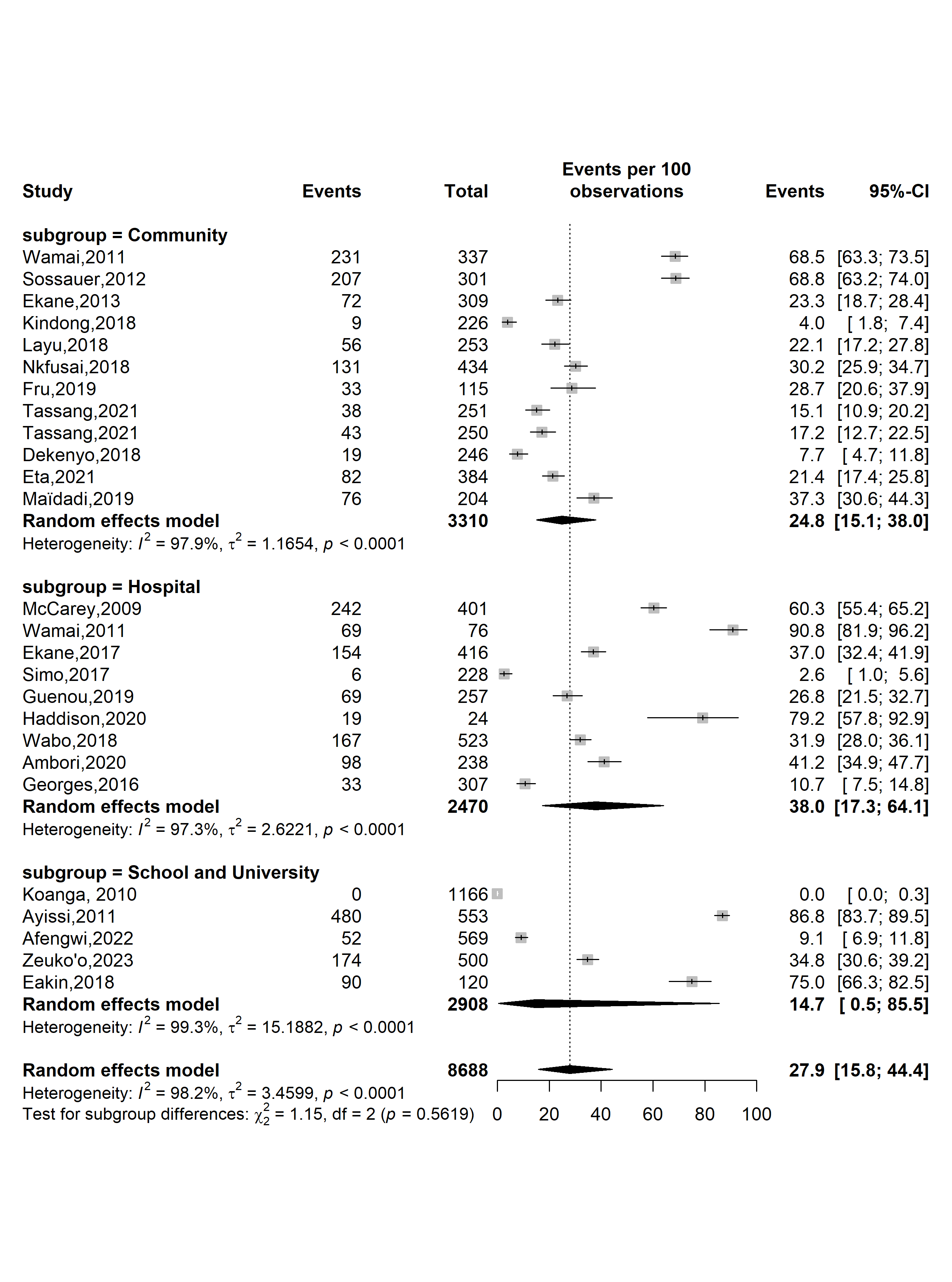

**Supplementary Fig. 2** Pooled prevalence of human papillomavirus knowledge as a cause of uterine cervical cancers by study settings in Cameroon

**Study site**

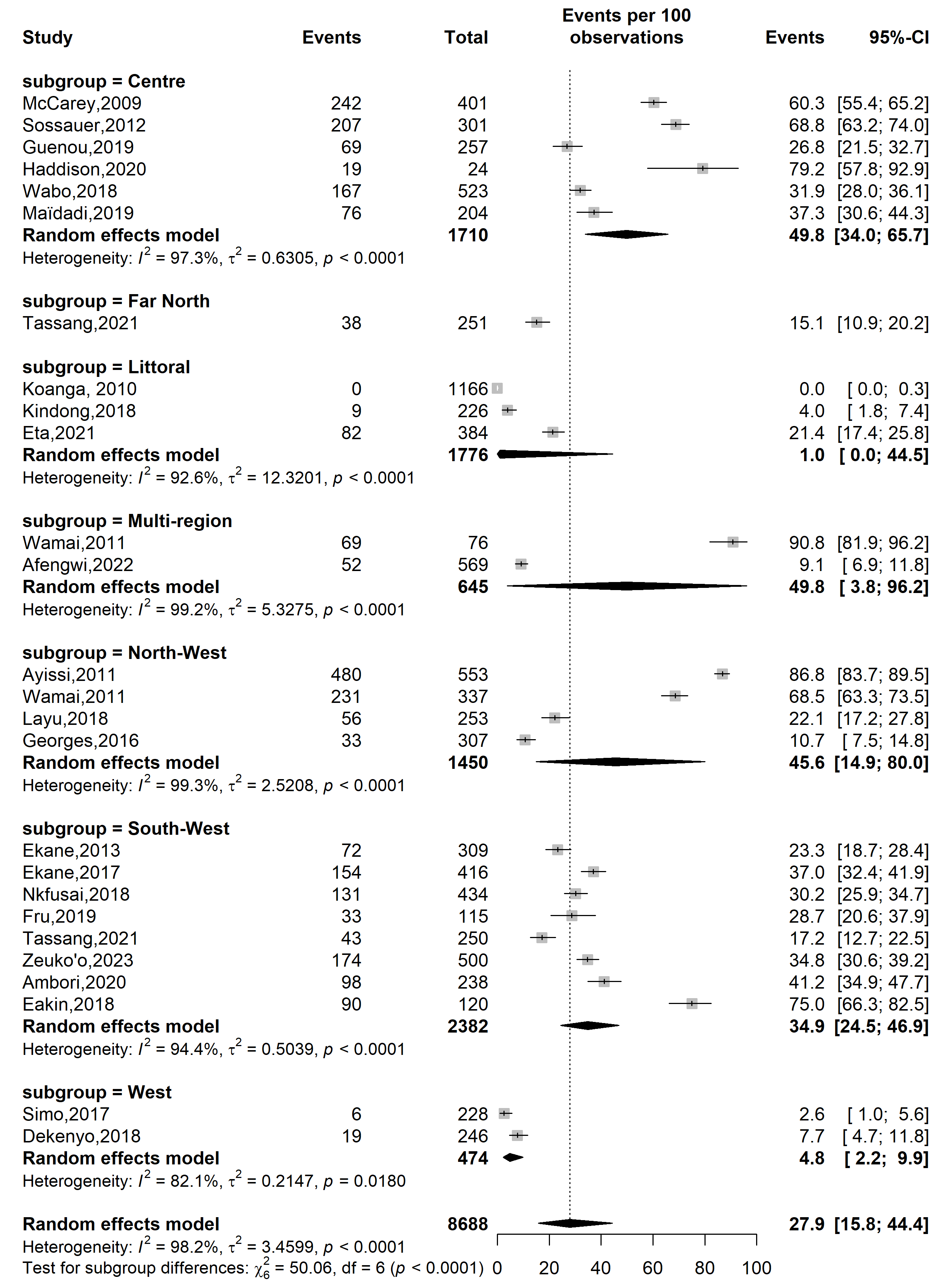

**Supplementary Fig. 3** Pooled prevalence of human papillomavirus knowledge as a cause of uterine cervical cancers by study sites in Cameroon

**Sampling method**

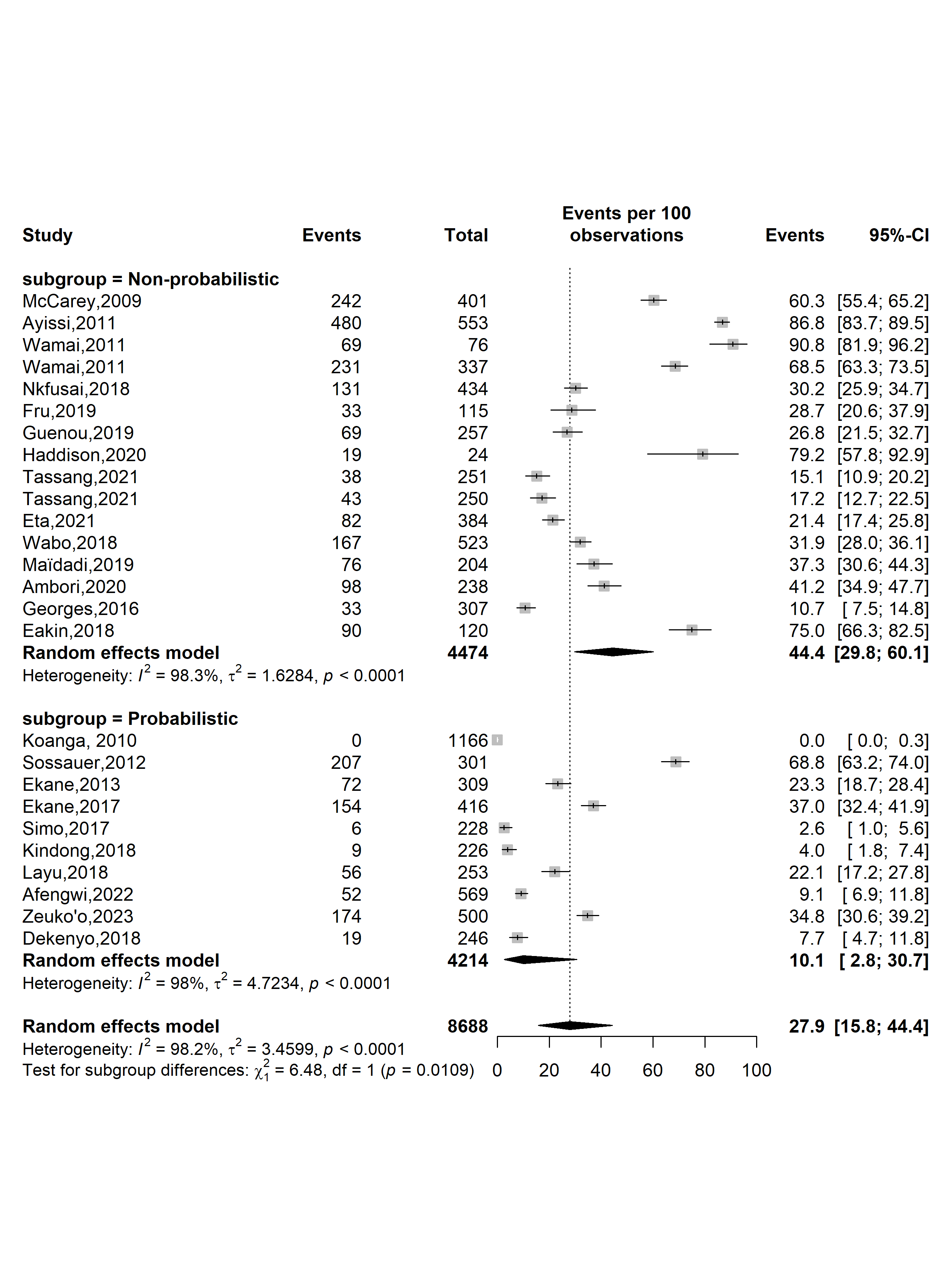

**Supplementary Fig.** Pooled prevalence of human papillomavirus knowledge as a cause of uterine cervical cancers by sampling methods in Cameroon

**Type of participants**

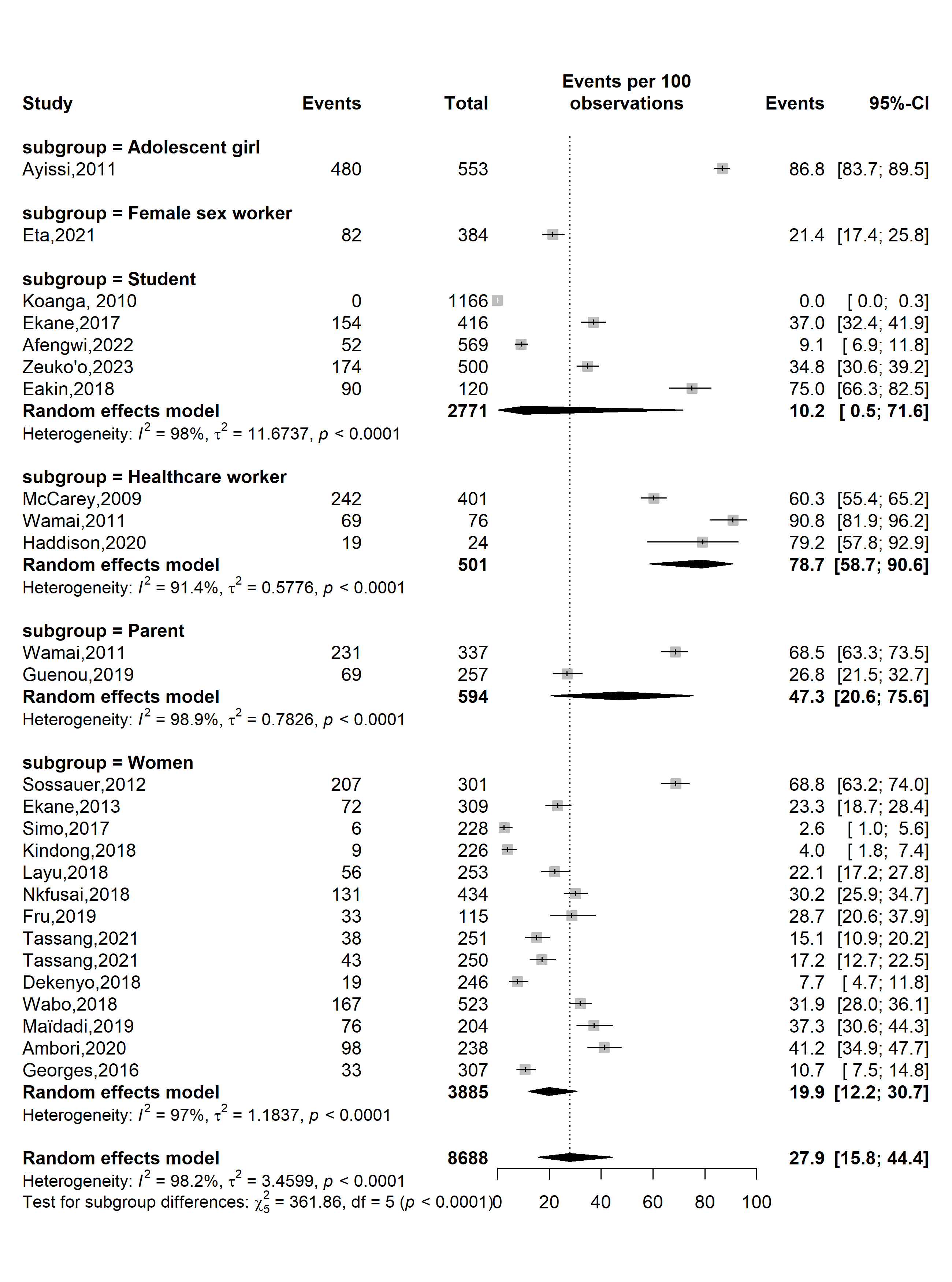

**Supplementary Fig. 5** Pooled prevalence of human papillomavirus knowledge as a cause of uterine cervical cancers by type of participants in Cameroon

**Sensitivity analysis**

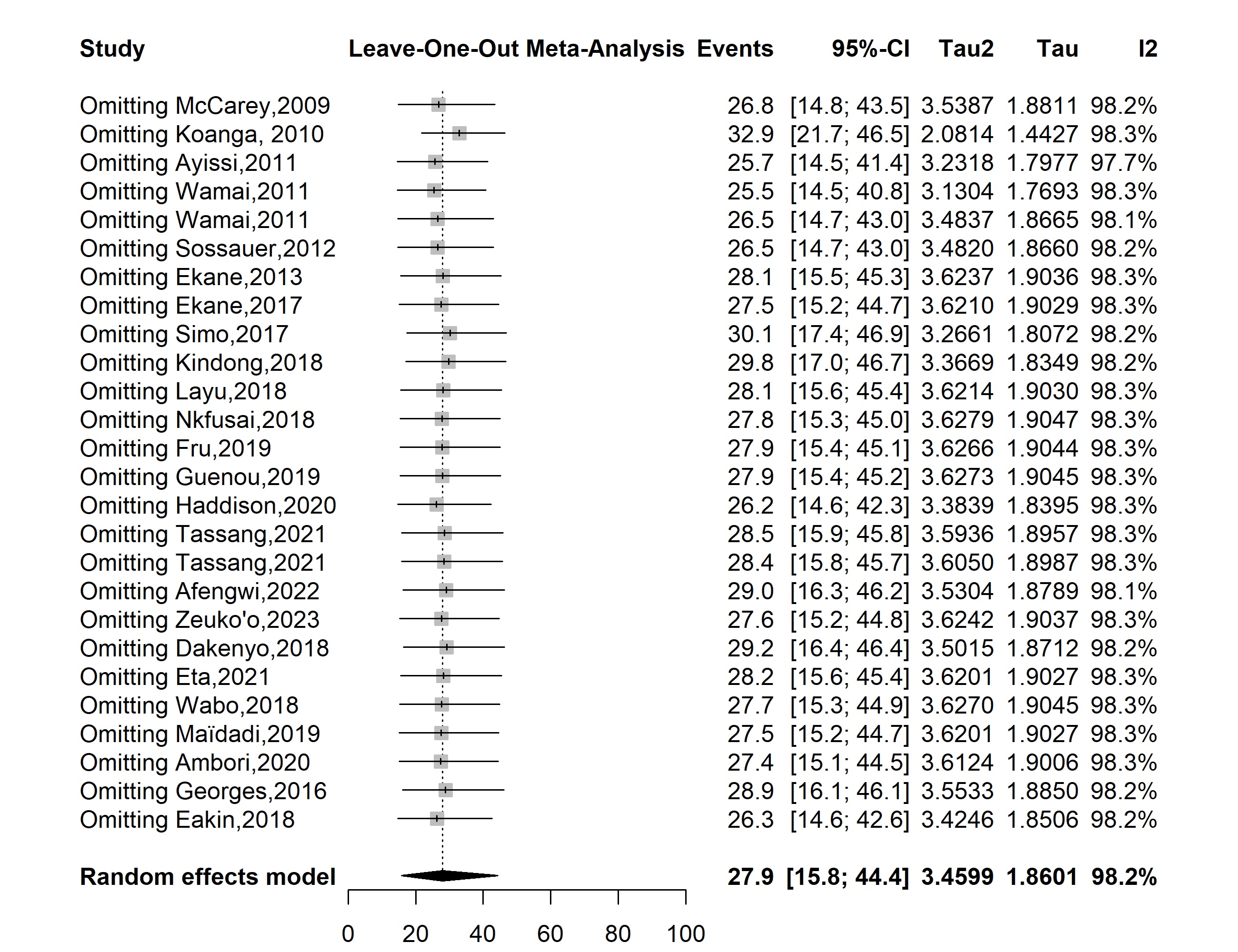

**Supplementary Fig. 6** Sensitivity analysis of the pooled prevalence of human papillomavirus knowledge as a cause of uterine cervical cancers in Cameroon

**Publication bias assessment**

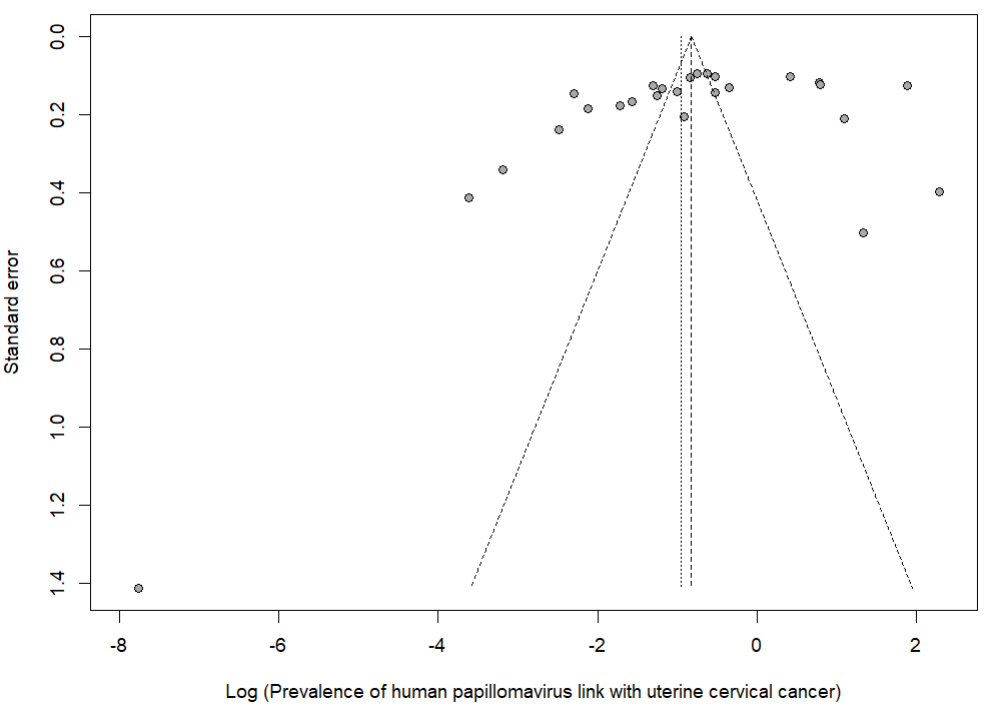

Egger’s test *p*-value = 0.213

Begg’s test *p*-value = 0.082

**Supplementary Fig. 7** Funnel plot displaying the risk of publication bias of studies assessing the prevalence of human papillomavirus knowledge as a cause of uterine cervical cancers in Cameroon
