## Supplementary Material 3 for "Human papillomavirus knowledge and associated factors in Cameroon: a systematic review and meta-analysis"

**Subgroup analysis**

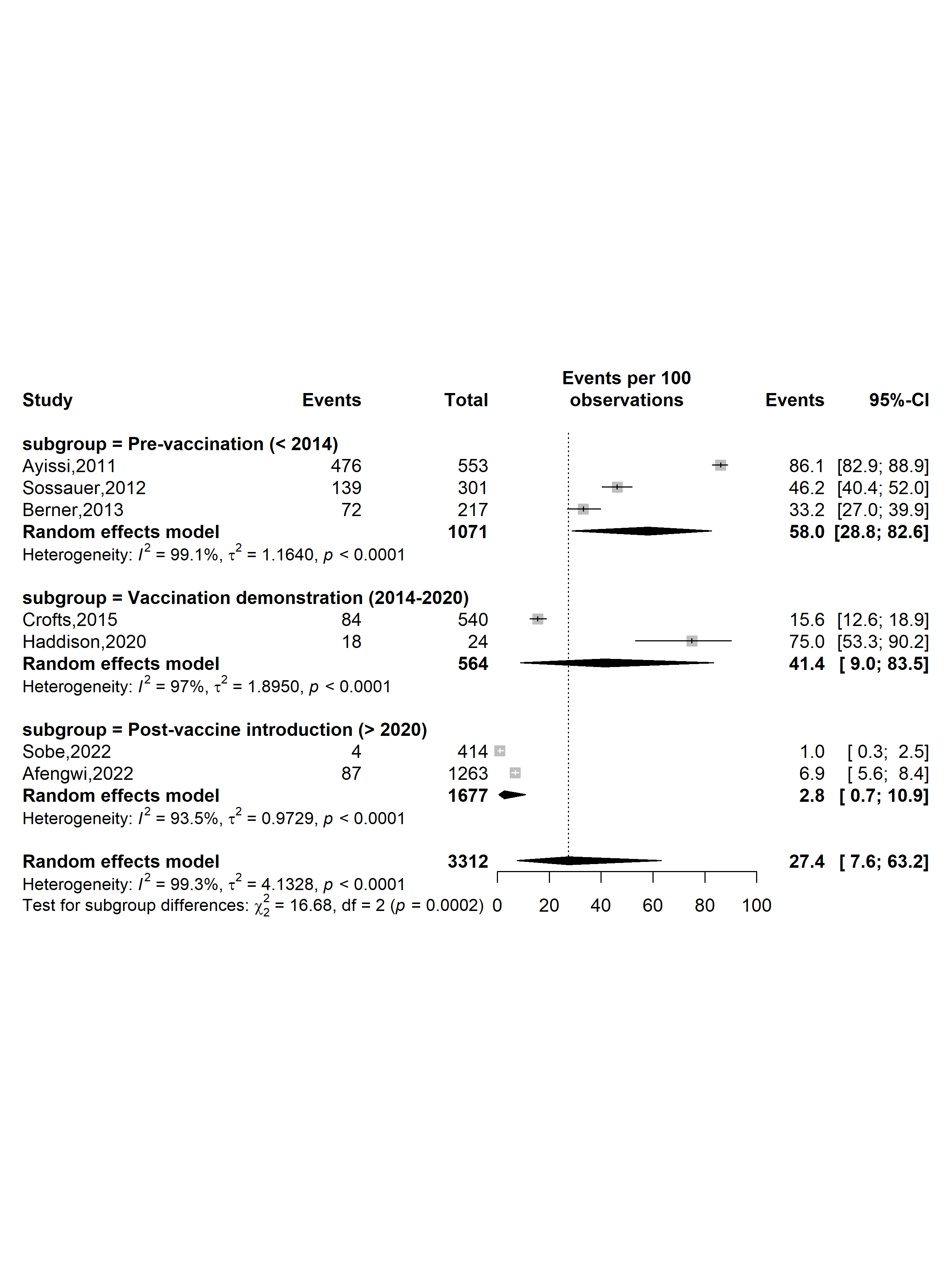

**Supplementary Fig. 1** Pooled prevalence of human papillomavirus (HPV) good knowledge by HPV vaccine introduction timeframe in Cameroon

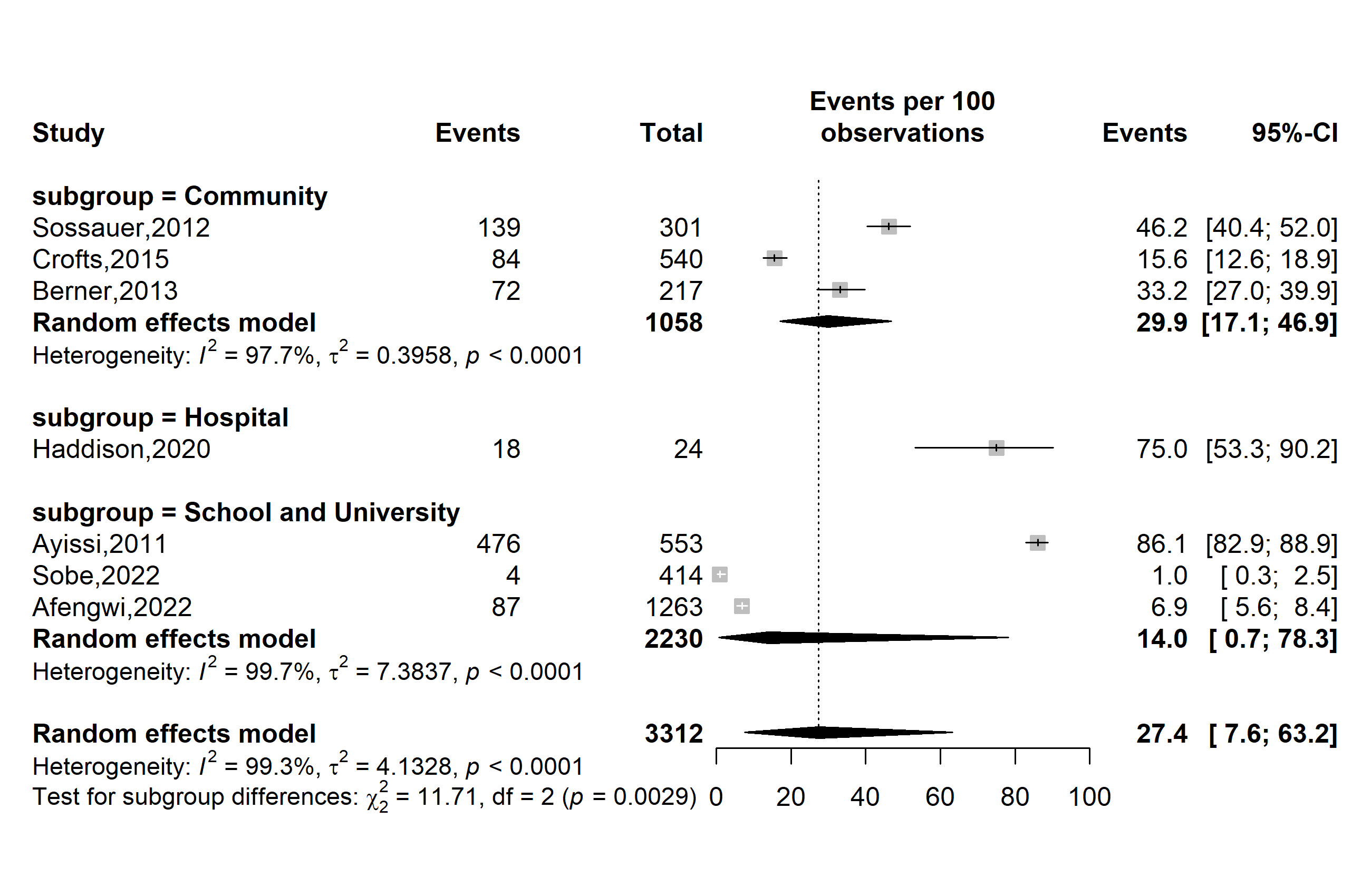

**Supplementary Fig. 2** Pooled prevalence of human papillomavirus (HPV) good knowledge by HPV vaccine introduction timeframe in Cameroon

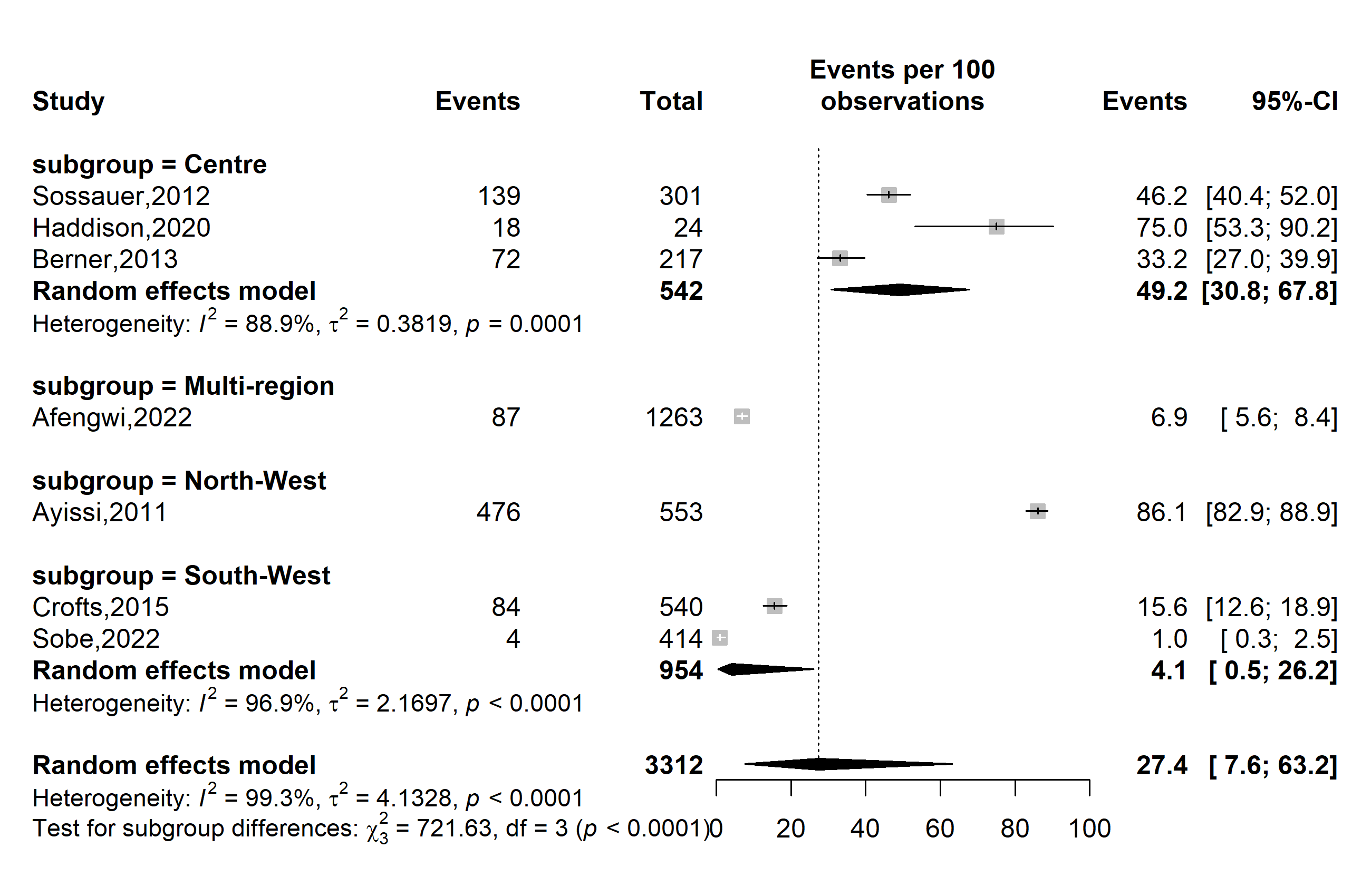

**Supplementary Fig. 3** Pooled prevalence of human papillomavirus (HPV) good knowledge by study site in Cameroon

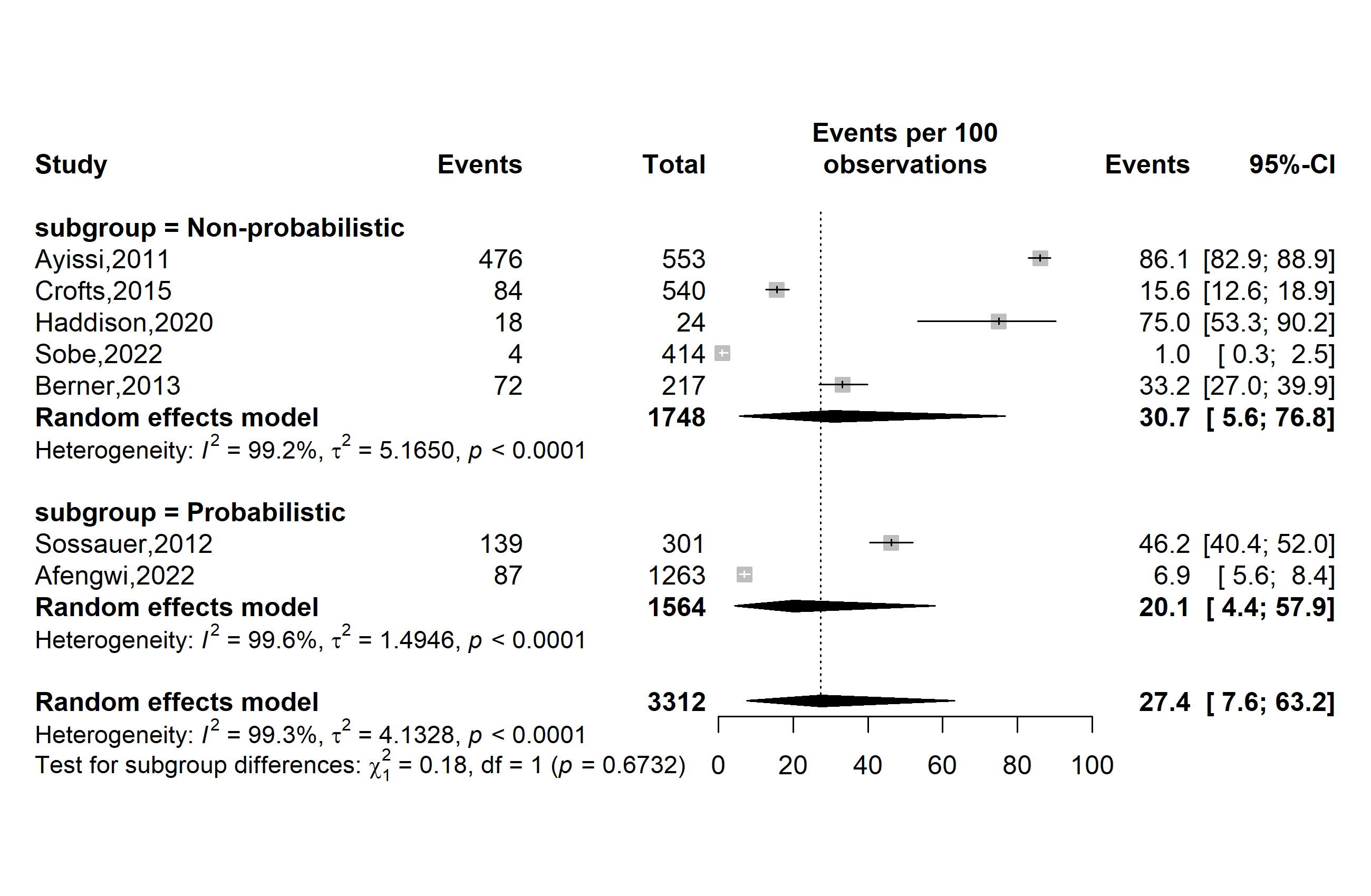

**Supplementary Fig. 4** Pooled prevalence of human papillomavirus (HPV) good knowledge by sampling method in Cameroon

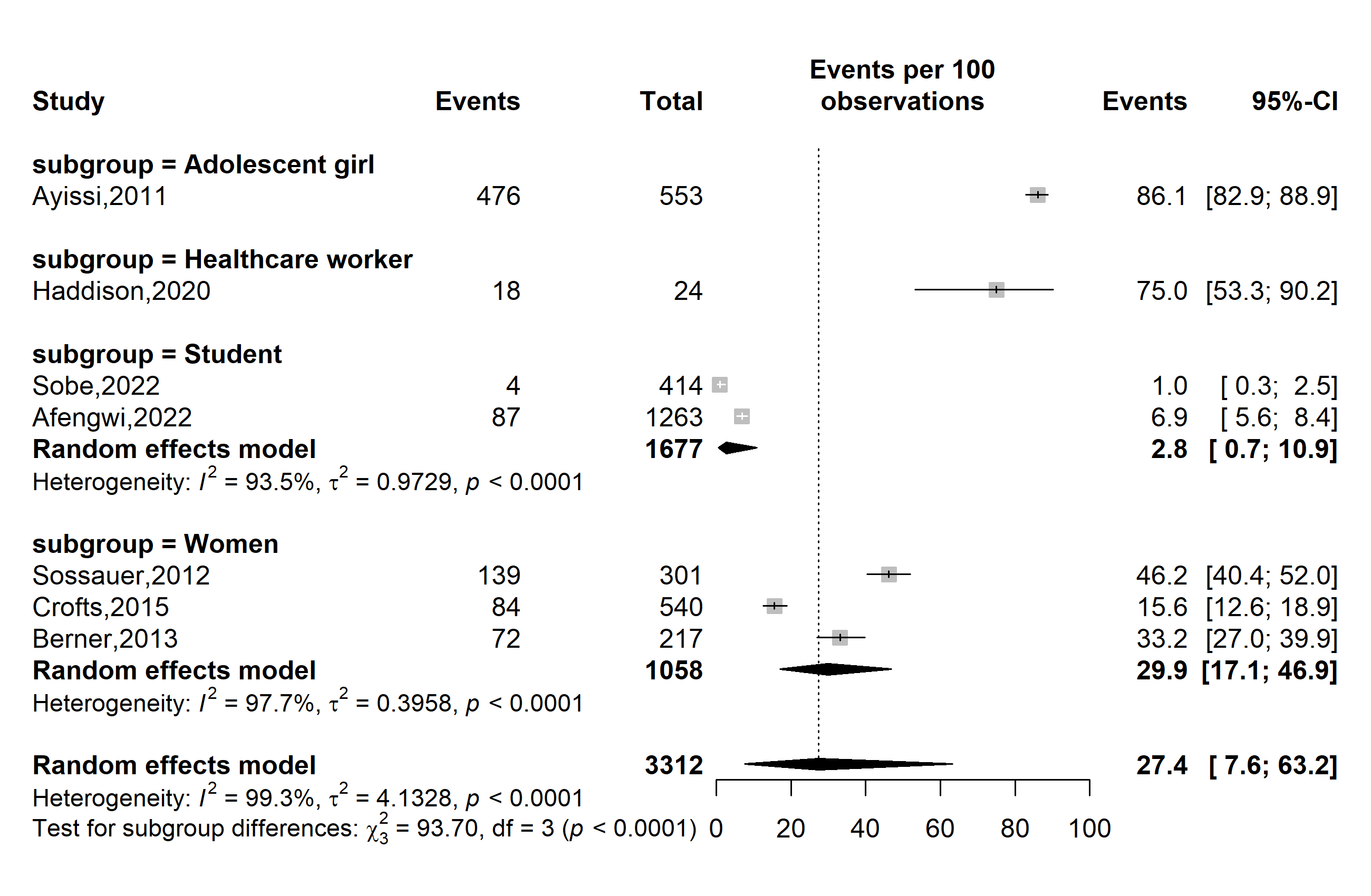

**Supplementary Fig. 5** Pooled prevalence of human papillomavirus (HPV) good knowledge by types of participants in Cameroon

**Sensitivity analysis**

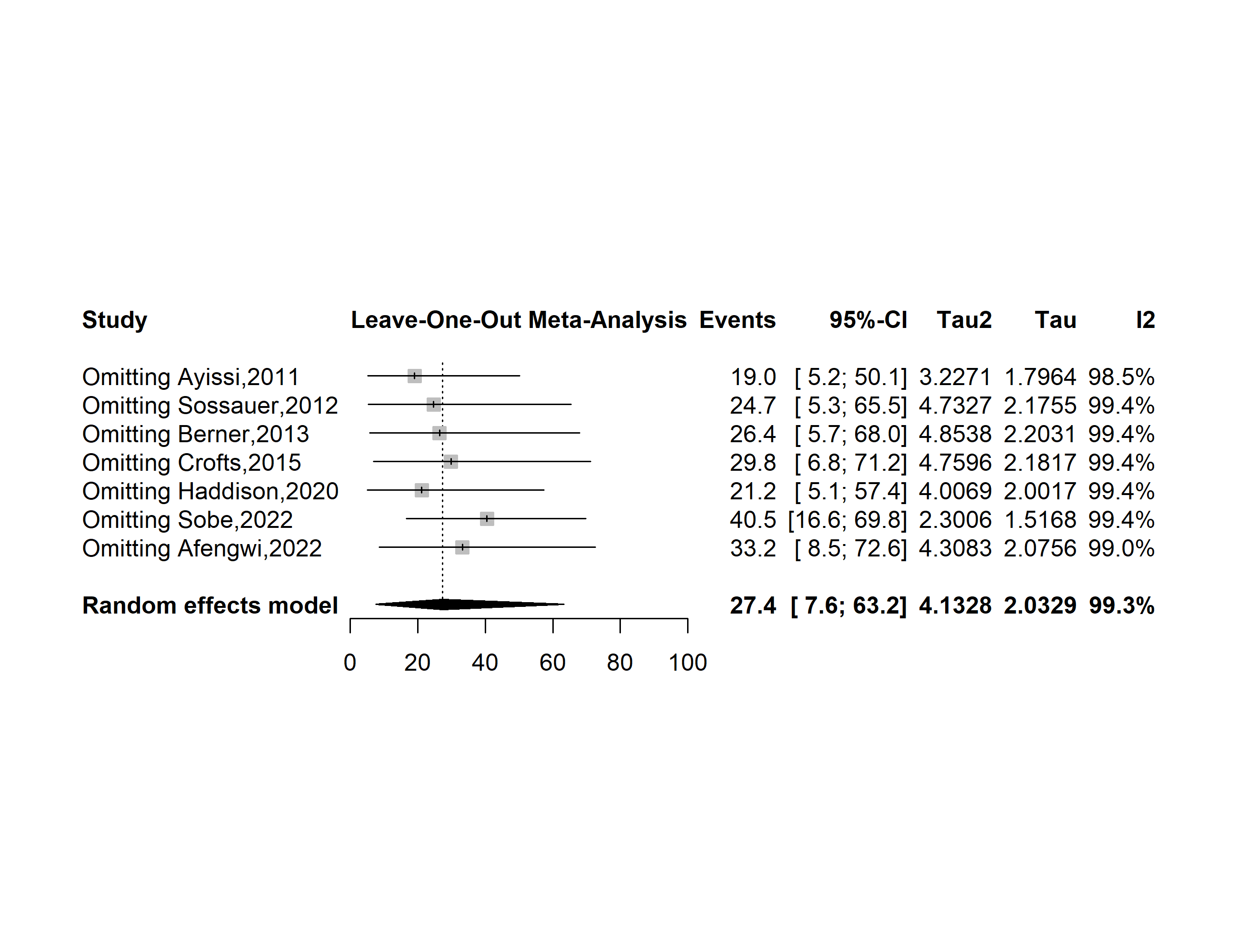

**Supplementary Fig. 6** Forest plot displaying the sensitivity analysis of pooled the prevalence of human papillomavirus good knowledge in Cameroon

**Publication bias assessment**

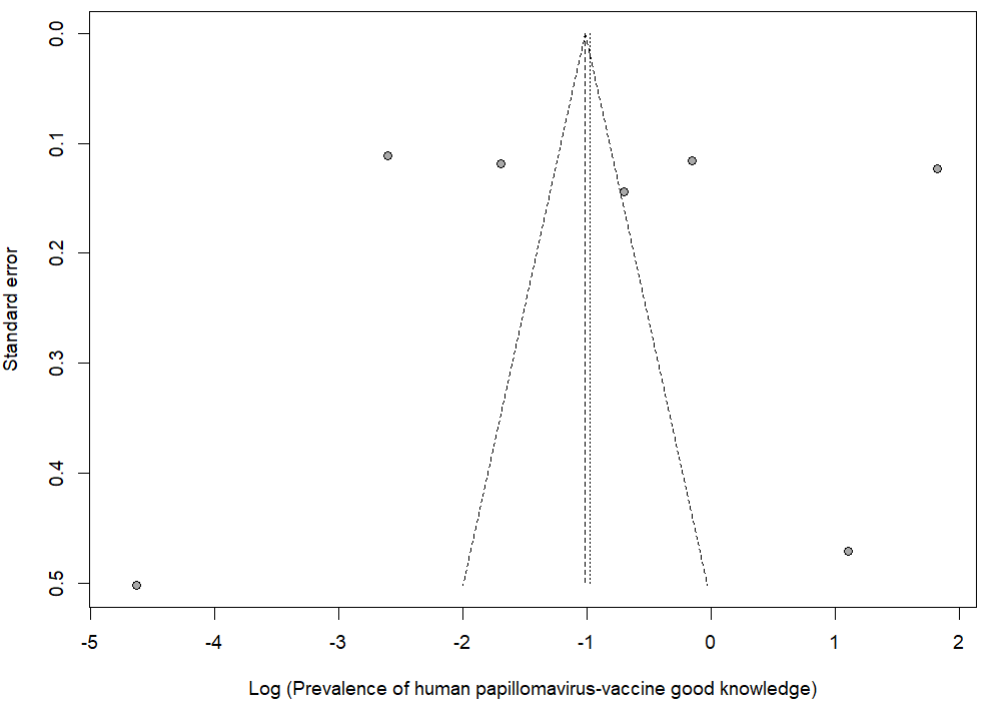

**Supplementary Fig. 7** Funnel plot displaying the risk of publication bias of studies assessing the prevalence of human papillomavirus good knowledge in Cameroon

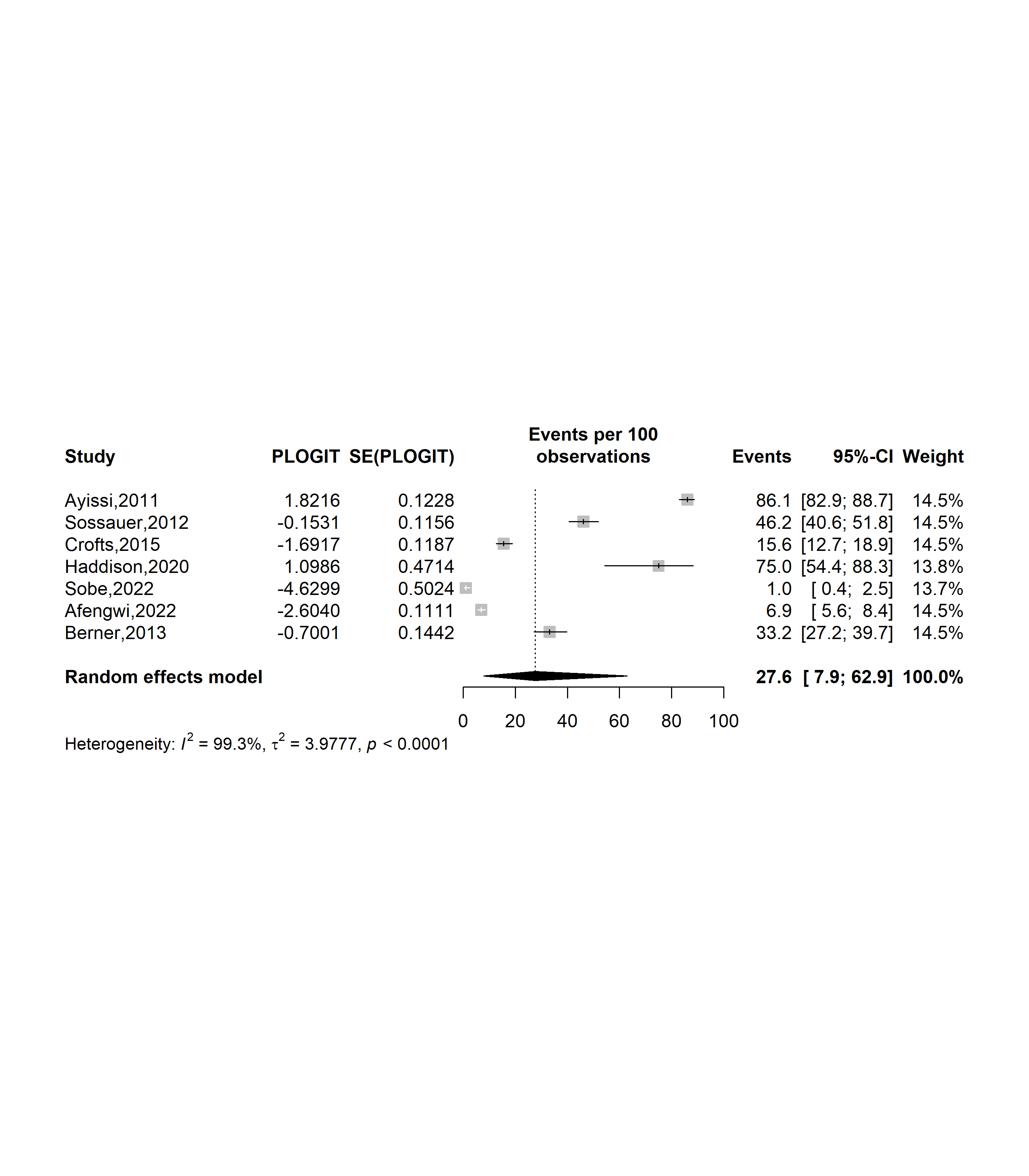

**Supplementary Fig. 8** Forest plot displaying the trim and fill analysis of studies assessing the prevalence of human papillomavirus good knowledge in Cameroon

**Risk factors**

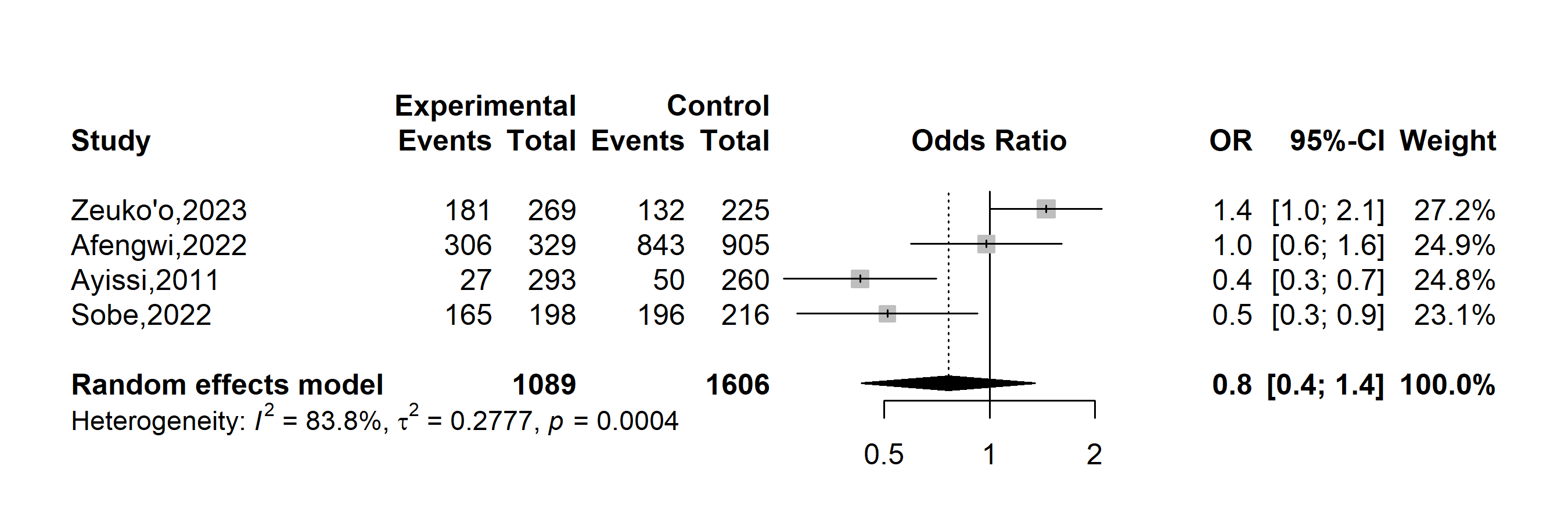

**Supplementary Fig. 9** Forest plot displaying pooled odds ratio of having poor knowledge related to human papillomavirus disease in Cameroon by age (˂ 20 vs. 20+)

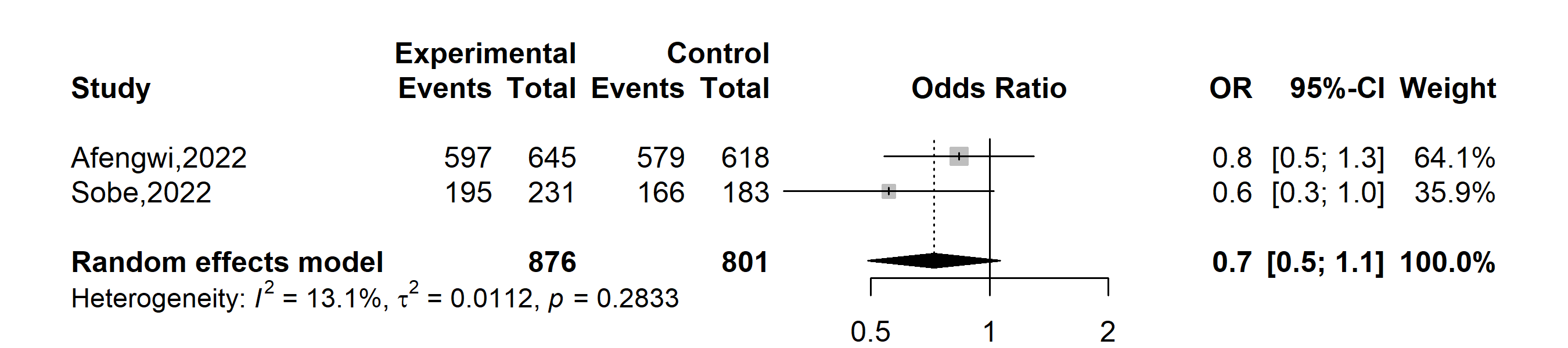

**Supplementary Fig. 10** Forest plot displaying pooled odds ratio of having poor knowledge related to human papillomavirus disease in Cameroon by gender (Female vs. Male)

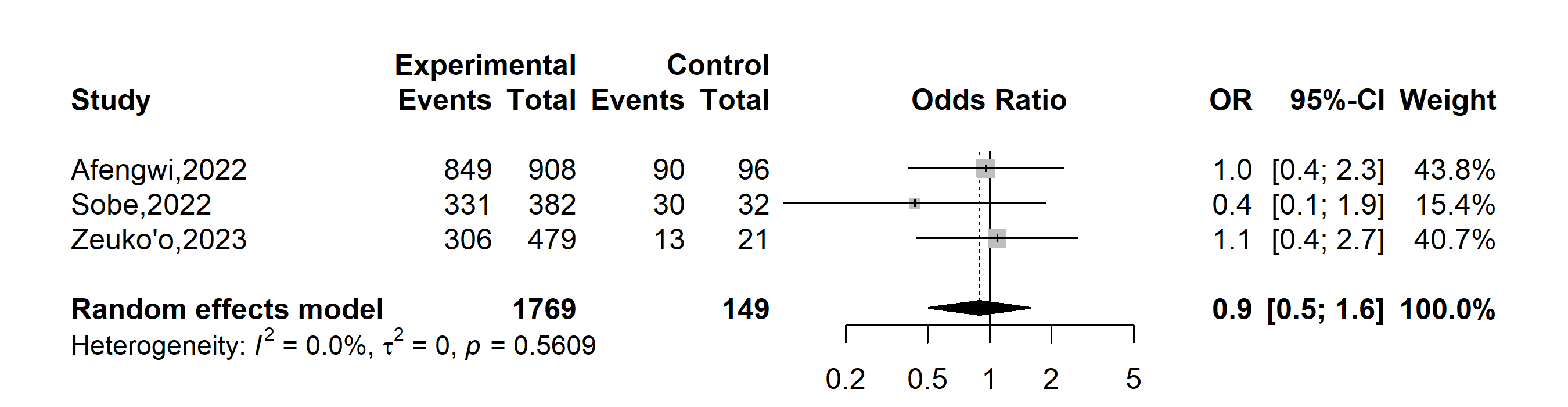

**Supplementary Fig. 11** Forest plot displaying pooled odds ratio of having poor knowledge related to human papillomavirus disease in Cameroon by marital status (Single vs. In partnership)

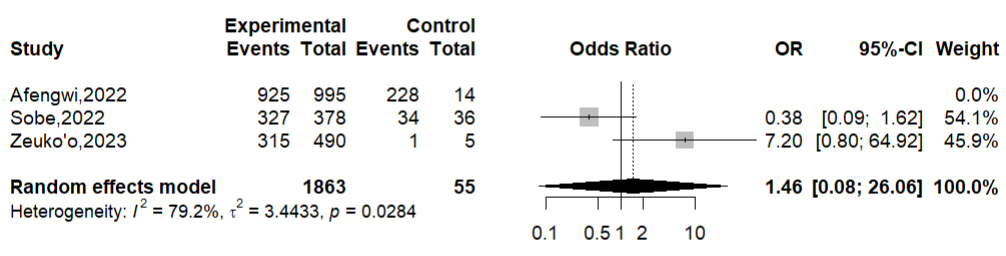

**Supplementary Fig. 12** Forest plot displaying pooled odds ratio of having poor knowledge related to human papillomavirus disease in Cameroon by religion (Christian vs. Others)

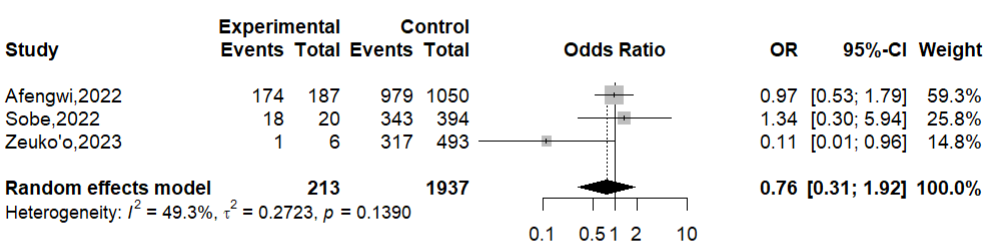

**Supplementary Fig. 13** Forest plot displaying pooled odds ratio of having poor knowledge related to human papillomavirus disease in Cameroon by religion (Muslim vs. Others)

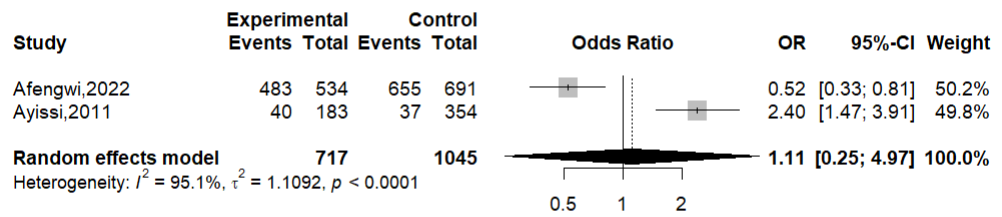

**Supplementary Fig. 14** Forest plot displaying pooled odds ratio of having poor knowledge related to human papillomavirus disease in Cameroon by educational level (Tertiary vs. Lower levels)

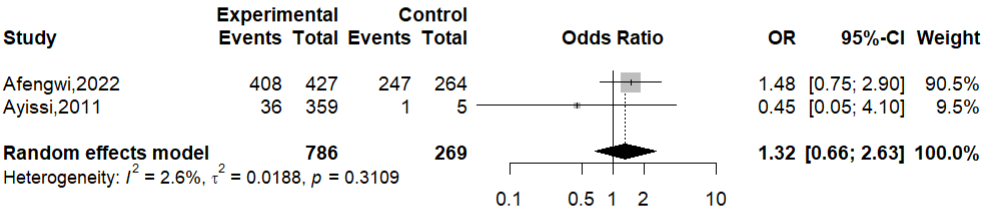

**Supplementary Fig. 14** Forest plot displaying pooled odds ratio of having poor knowledge related to human papillomavirus disease in Cameroon by educational level (Secondary vs. Primary)
