## Supplementary Material 1 for "Human papillomavirus knowledge and associated factors in Cameroon: a systematic review and meta-analysis"

**Supplementary Table 1** Searching strategy by database

| **Database** | **Search string** | **Number of entries** |
| --- | --- | --- |
| **PubMed** | ("human papillomavirus" OR "HPV" OR "cervical cancer") AND ("knowledge" OR “attitude” OR “awareness” OR “practice”) AND ("Cameroon" OR "Cameroonian" OR "Cameroun") | 60 |
| **Scopus** | TITLE-ABS-KEY ("human papillomavirus" OR "hpv" OR “cervical cancer")  AND  TITLE-ABS-KEY ("knowledge" OR “attitude” OR “awareness” OR “practice”) AND  TITLE-ABS-KEY ("Cameroon" OR "Cameroonian" OR "Cameroun") | 55 |
| **Web of sciences** | ("human papillomavirus" OR "HPV" OR "cervical cancer")  AND ("knowledge" OR "awareness" OR "attitude" OR "practice")  AND  ("Cameroon" OR "Cameroonian" OR "Cameroun") | 134 |
| **Embase** | ("human papillomavirus" OR "HPV" OR "cervical cancer")  AND ("knowledge" OR "awareness" OR "attitude" OR "practice")  AND  ("Cameroon" OR "Cameroonian" OR "Cameroun") | 72 |
| **Cochrane Library** | (“human papillomavirus” OR HPV OR “cervical cancer”)  AND  (knowledge OR awareness OR attitude OR practice)  AND  (Cameroon OR Cameroonian OR Cameroun) | 8 |
| **African Journals Online (AJOL)** | (“human papillomavirus” OR HPV OR “cervical cancer”)  AND  (vaccine OR vaccination OR knowledge OR awareness OR attitude OR practice)  AND  (Cameroon OR Cameroonian OR Cameroun) | 77 |
| **Health Sciences and Disease** | (“human papillomavirus” OR HPV OR “cervical cancer”)  AND  (knowledge OR awareness OR attitude OR practice)  AND  (Cameroon OR Cameroonian OR Cameroun) | 1 |

**Subgroup analysis**

**Study period**

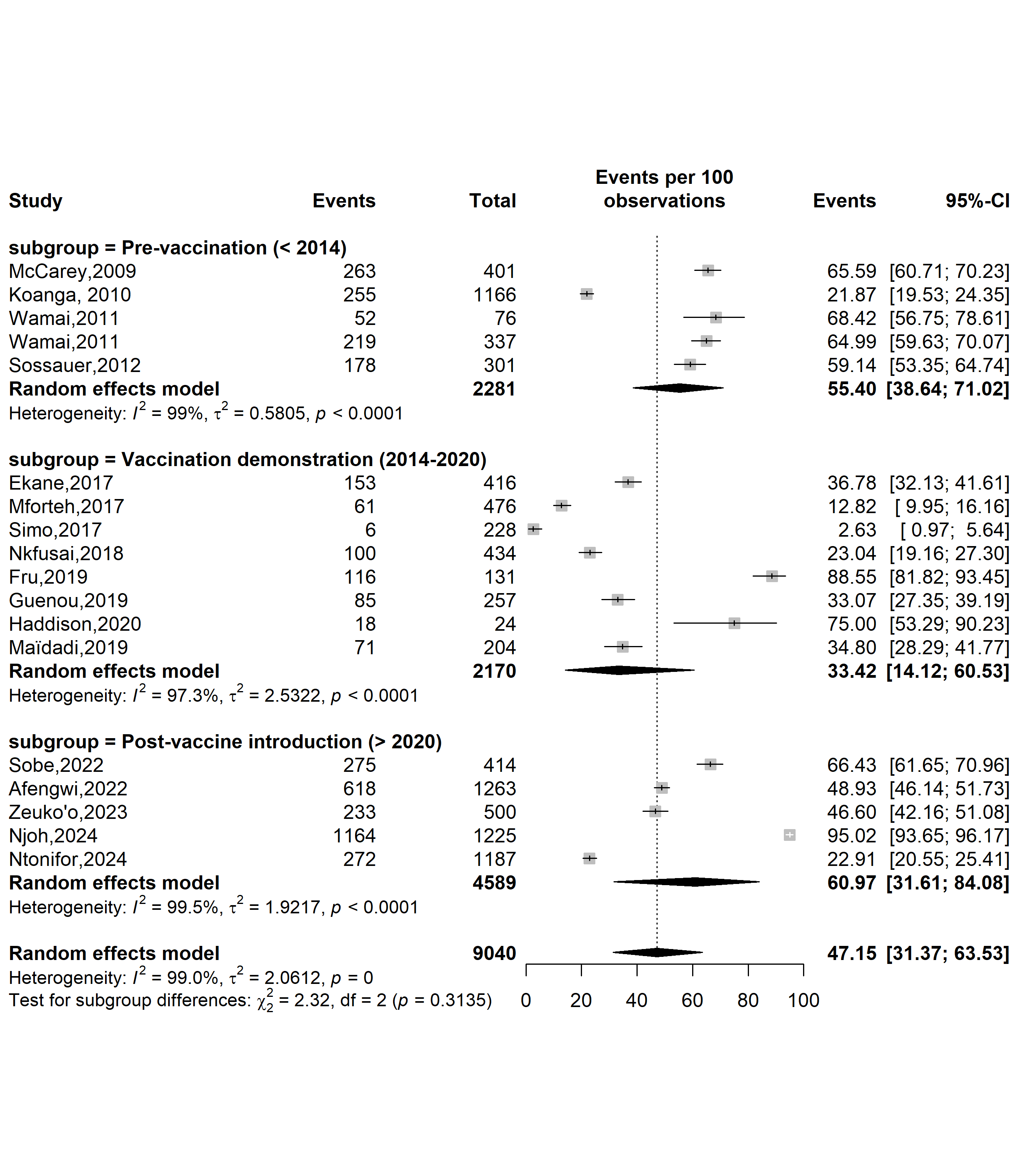

**Supplementary Fig. 1** Pooled prevalence of human papillomavirus knowledge (HPV) as a sexually transmitted disease by HPV vaccine introduction timeframe in Cameroon

**Study setting**

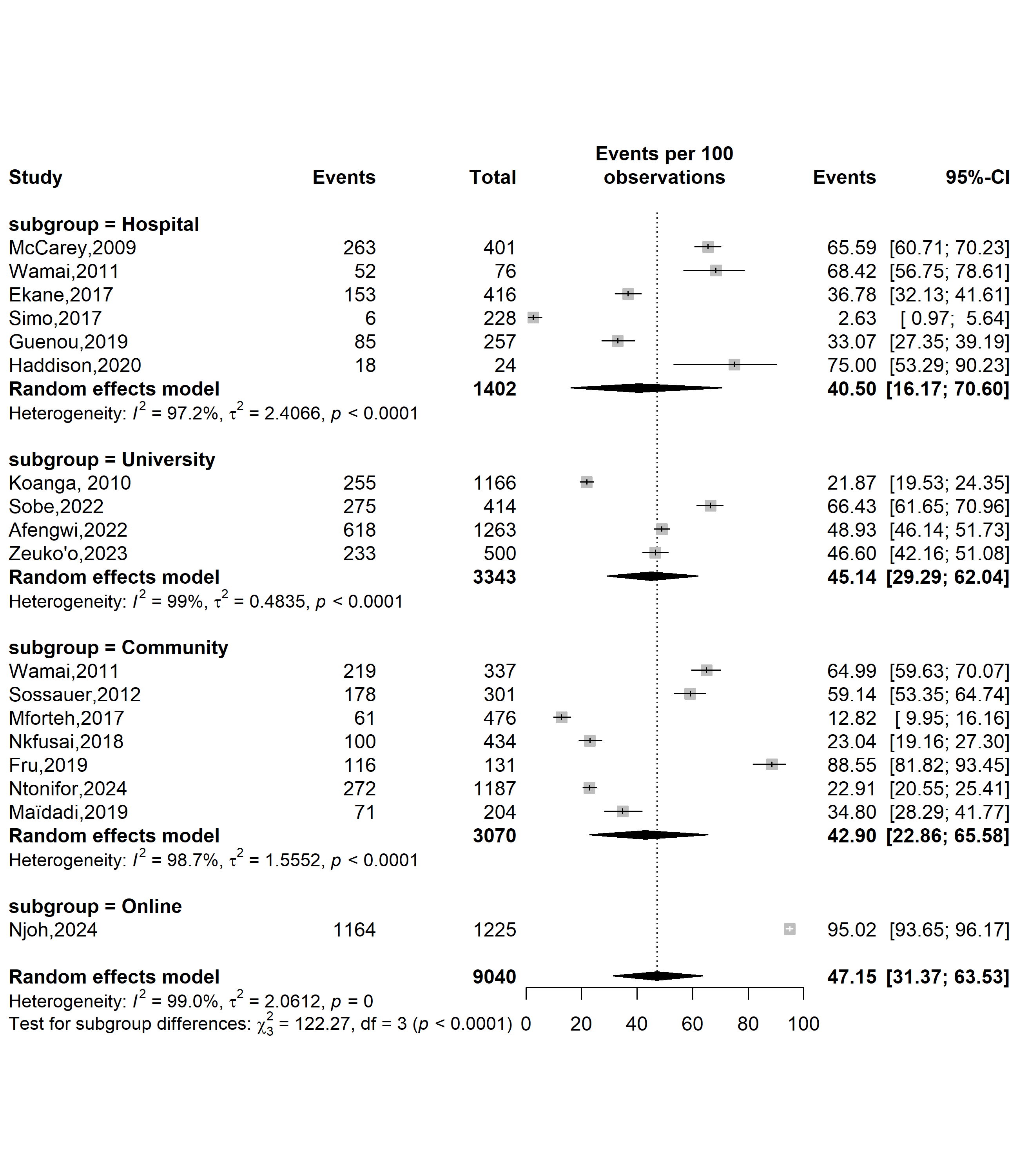

**Supplementary Fig. 2** Pooled prevalence of human papillomavirus knowledge as a sexually transmitted disease by study settings in Cameroon

**Study site**

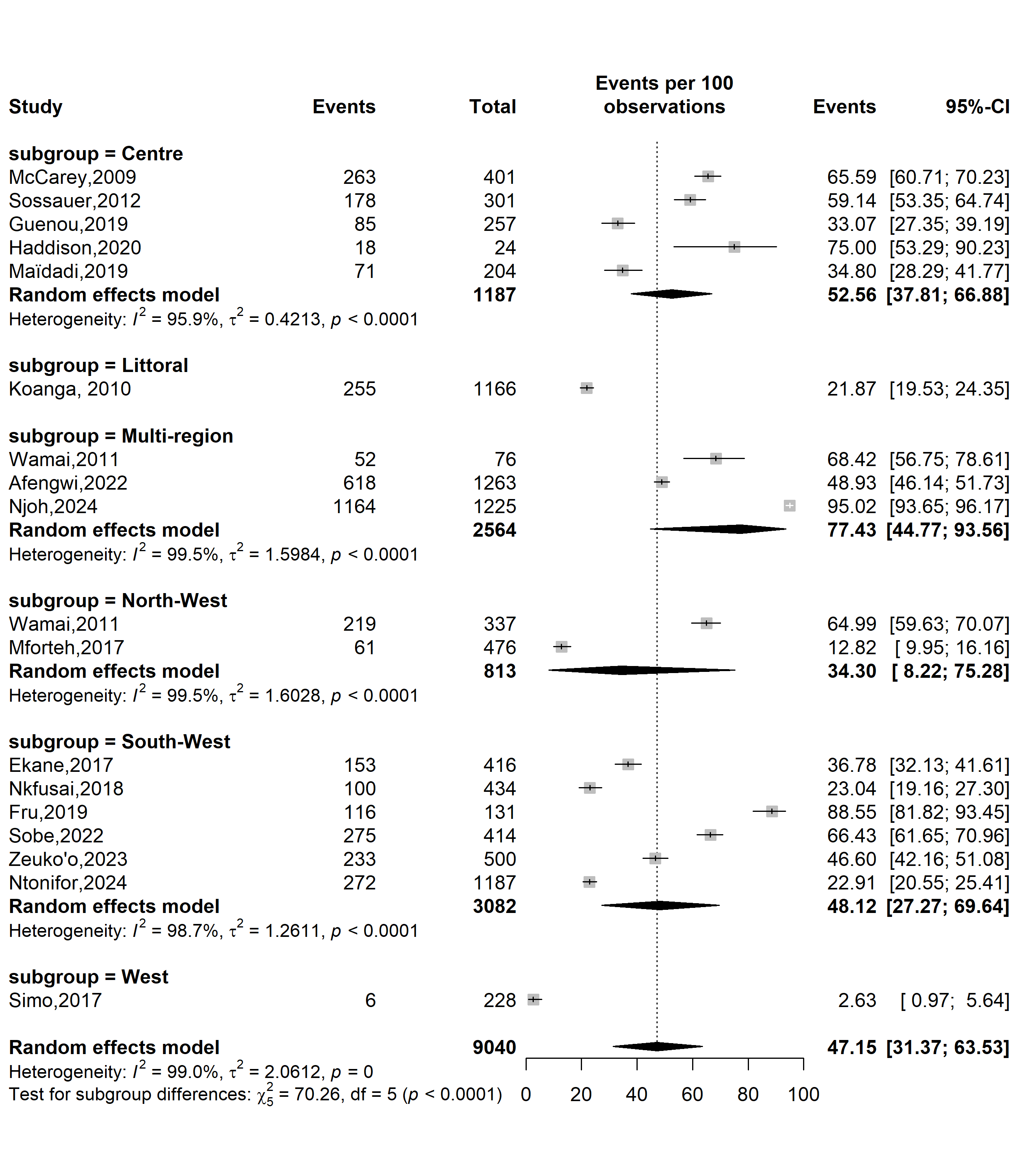

**Supplementary Fig. 3** Pooled prevalence of human papillomavirus knowledge as a sexually transmitted disease by study sites in Cameroon

**Sampling method**

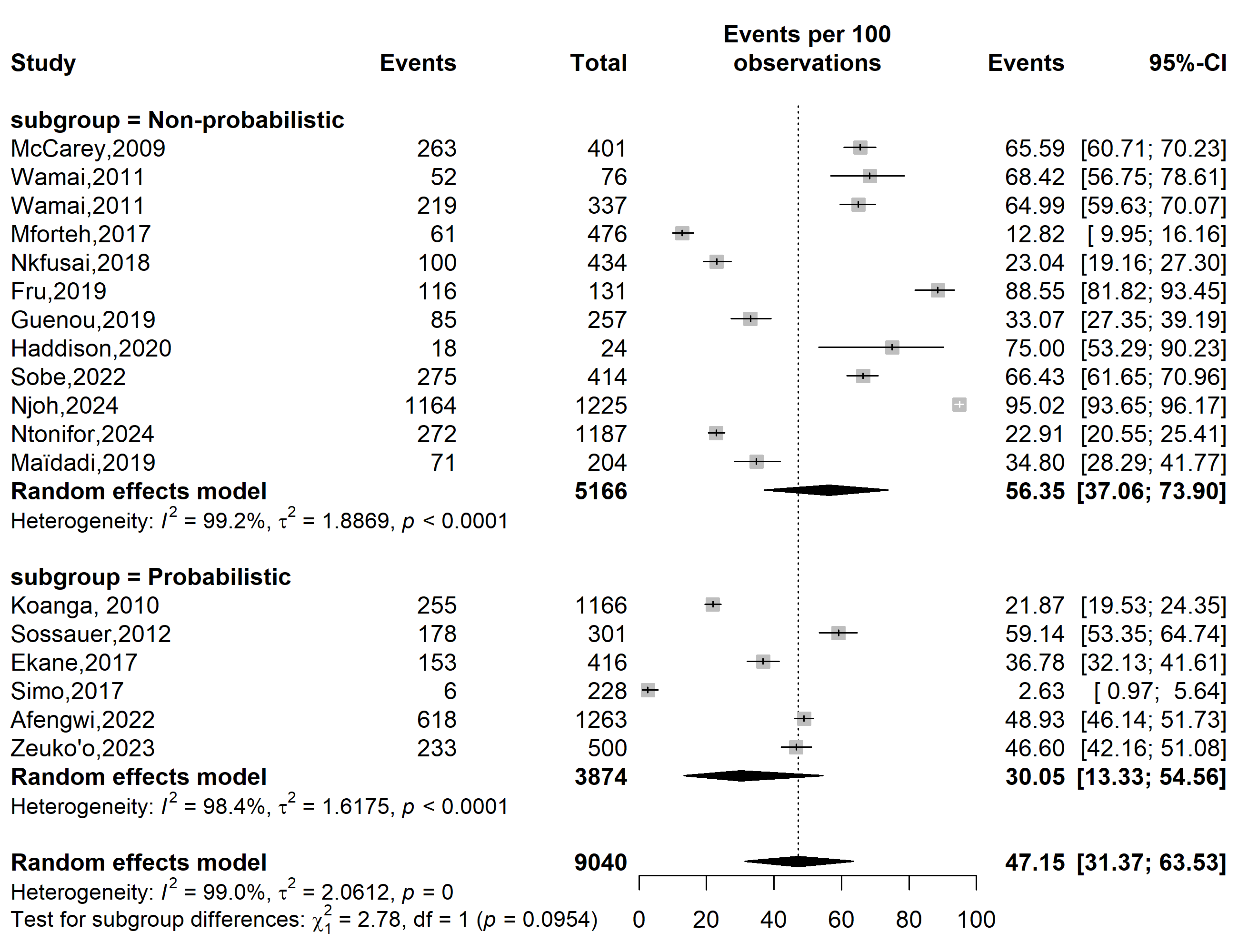

**Supplementary Fig. 4** Pooled prevalence of human papillomavirus knowledge as a sexually transmitted disease by sampling methods in Cameroon

**Type of participants**

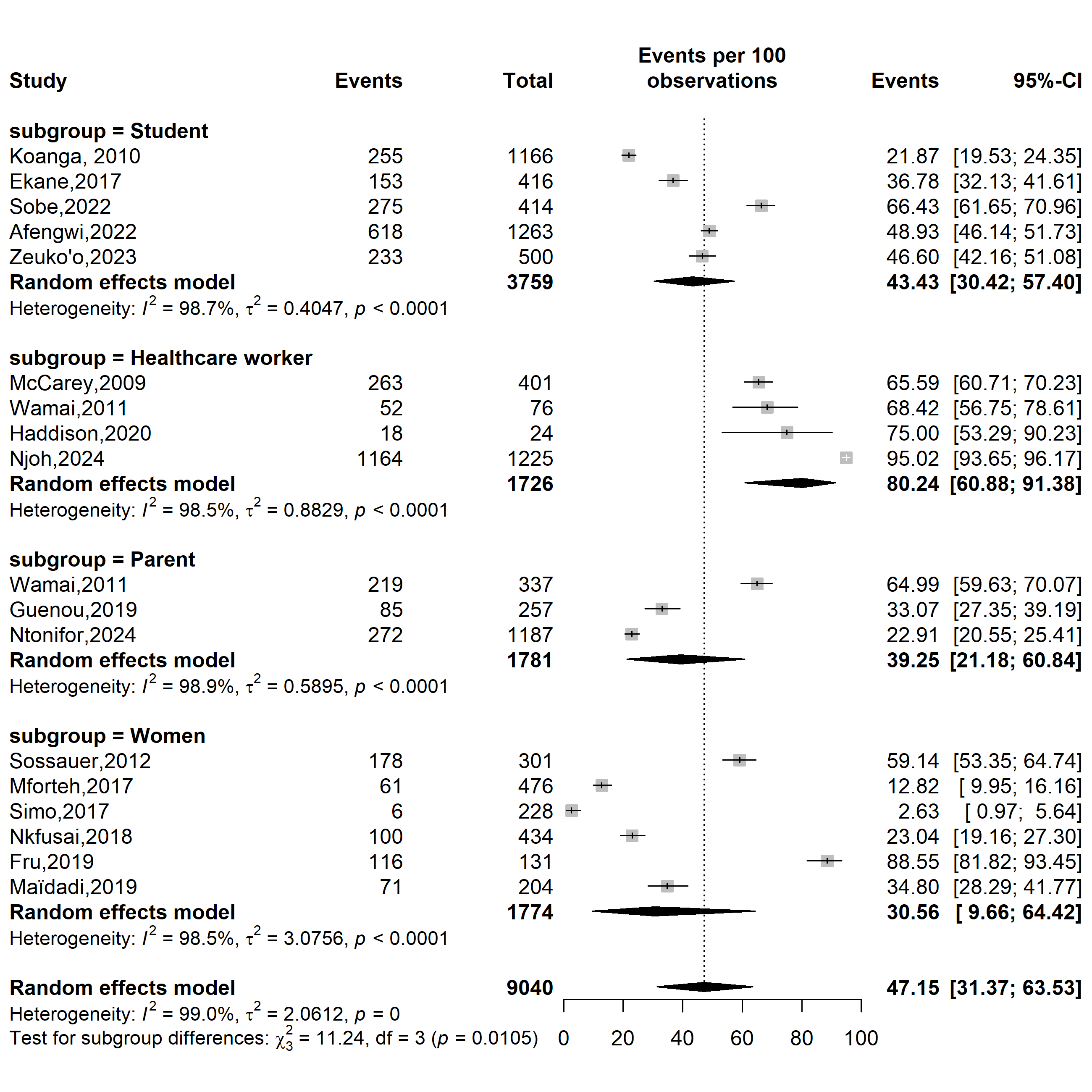

**Supplementary Fig. 5** Pooled prevalence of human papillomavirus knowledge as a sexually transmitted disease by types of participants in Cameroon

**Sensitivity analysis**

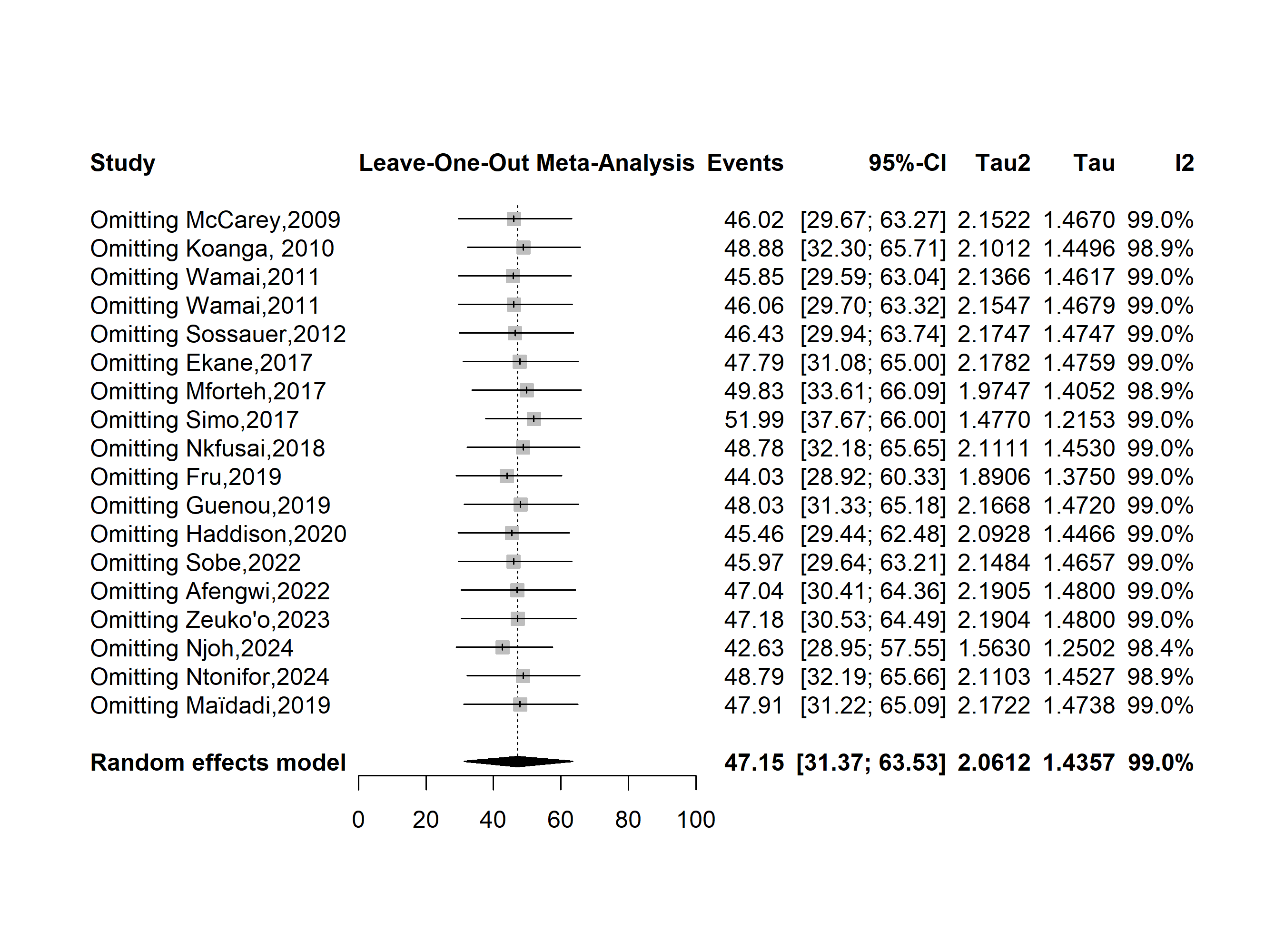

**Supplementary Fig. 6** Sensitivity analysis of human papillomavirus knowledge as a sexually transmitted disease in Cameroon

**Publication bias assessment**

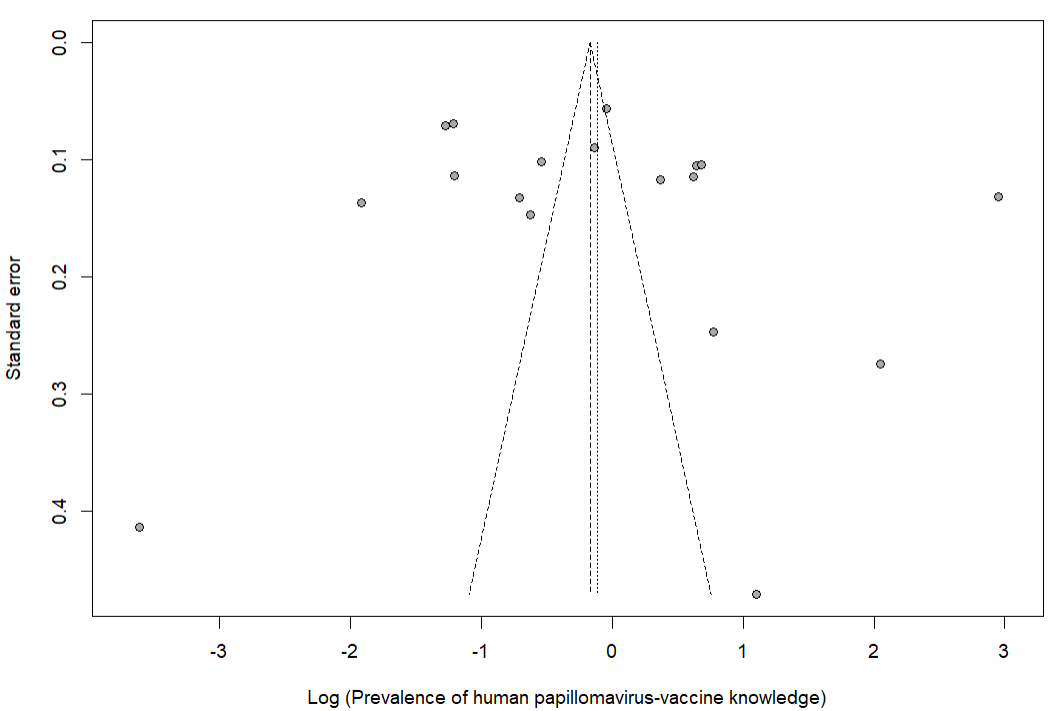

Egger’s test *p*-value = 0.414

Begg’s test *p*-value = 0.677

**Supplementary Fig. 7** Funnel plot displaying the risk of publication bias of studies assessing human papillomavirus knowledge as a sexually transmitted disease in Cameroon
